# Population impact of SARS-CoV-2 variants with enhanced transmissibility and/or partial immune escape

**DOI:** 10.1101/2021.08.26.21262579

**Authors:** Mary Bushman, Rebecca Kahn, Bradford P. Taylor, Marc Lipsitch, William P. Hanage

## Abstract

SARS-CoV-2 variants of concern exhibit varying degrees of transmissibility and, in some cases, escape from infection- and vaccine-induced immunity. Much effort has been devoted to measuring these phenotypes, but predicting their impact on the course of the pandemic – especially that of immune escape – remains a challenge. Here, we use a mathematical model to simulate the dynamics of wildtype and variant strains of SARS-CoV-2 in the context of vaccine rollout and nonpharmaceutical interventions. We show that variants with enhanced transmissibility easily rise to high frequency, whereas partial immune escape, on its own, often fails to do so. However, when these phenotypes are combined, enhanced transmissibility can carry the variant to high frequency, at which point partial immune escape may limit the ability of vaccination to control the epidemic. Our findings suggest that moderate immune escape poses a low risk unless combined with a substantial increase in transmissibility.

## Introduction

The second year of the COVID-19 pandemic has been dominated by variants of concern – SARS-CoV-2 lineages that have driven resurgent waves of the disease, often more severe than earlier waves. The World Health Organization currently recognizes four variants of concern (VOCs): Alpha (previously designated B.1.1.7), which was first identified in the United Kingdom; Beta (B.1.351), first reported in South Africa; Gamma (P.1), believed to have originated in Brazil; and Delta (B.1.617), first detected in India. Initially reported in late 2020 or early 2021, these four variants are now found on every continent except Antarctica [1].

The speed and consistency with which these variants invade have been attributed to some combination of enhanced transmissibility and partial immune escape. Alpha is estimated to be 43-100% more transmissible than wildtype [2, 3], but is similarly neutralized by convalescent sera [4-6] and is not associated with increased risk of reinfection [7]. There is some uncertainty regarding the transmissibility and immune escape of Beta because current data do not identify these individually, but result in estimates of these quantities that are inversely correlated. If immune escape is minimal, Beta may be up to 50% more transmissible than WT [8]. However, there is considerable evidence suggesting a moderate degree of immune escape, with significantly reduced neutralization by convalescent sera [4, 6, 9-11]; multiple studies have also reported that 40-50% of convalescent serum samples exhibit no neutralizing activity against Beta virus or pseudovirus [4, 10, 11]. (However, T cell responses may remain largely intact even when antibody responses are compromised [12, 13]). Gamma is believed to have significantly increased transmissibility, in the range of 70-140% higher than WT [14], and perhaps some degree of immune escape, with a modest reduction in neutralization by convalescent sera [15]. Gamma may reduce protection against reinfection by 21-46%, although as with Beta, estimates of transmissibility and immune escape are correlated [14]. Finally, evidence indicates that Delta is highly transmissible, with multiple analyses suggesting a 60% or greater increase in transmissibility relative to Alpha [16, 17]. Preliminary results suggest moderately reduced neutralization by convalescent serum [18, 19] as well as modest reductions in vaccine effectiveness compared to Alpha [20].

The appearance of these variants has taken place against a backdrop of accelerated vaccine development and rollout. Numerous candidate COVID-19 vaccines entered Phase III trials in the latter half of 2020, and many began distribution in late 2020 or early 2021. As of August 23, nearly 5 billion vaccine doses have been administered, but coverage is highly variable: 15 countries have given at least one dose to over three-quarters of their populations, while others have vaccinated less than one per hundred [21]. Most authorized vaccines demonstrate at least 70% efficacy against symptomatic COVID-19, and a few approach or exceed 90% efficacy [22-27]. Instances of severe disease are relatively rare in vaccinated individuals, and deaths exceedingly rare, so estimates of efficacy against severe outcomes tend to be imprecise, but true efficacy against severe disease and death is likely very high for most vaccines.

Along with evidence for partial escape from naturally acquired immunity, there is mounting evidence that vaccines may offer reduced protection against some variants. There is little evidence to suggest that Alpha evades vaccine-induced immunity; neutralization by post-vaccination sera is similar or slightly reduced compared to WT [4-6, 12, 28, 29], and minimal differences in vaccine efficacy have been observed [23, 30, 31]. However, numerous studies report markedly reduced neutralization of Beta by post-vaccination sera [4, 11, 12, 32-34], and some observational studies suggest moderately lower vaccine efficacy against Beta [24, 31, 35], although others show no reduction [36]. There is evidence suggesting slightly reduced neutralization of both Gamma and Delta by post-vaccination sera [15, 18, 19, 34, 37, 38]; in the case of Delta, vaccine efficacy may be slightly reduced as well [20].

For individuals, the implications of variants with partial vaccine escape are straightforward: protection decreases linearly with the product of the variant’s frequency in the population and the reduced efficacy against that variant. The risks to an entire population are much less clear. Due to the nonlinearity of epidemiological dynamics, population outcomes do not simply mirror those of individuals; a 30% reduction in vaccine efficacy does not translate to 30% fewer infections prevented, or 30% more infections overall. The same goes for transmissibility: a 50% increase in the average number of secondary infections from each case does not simply increase the total number of infections by 50%.

The difficulty of projecting population-level outcomes from variant phenotypes has limited our ability to distinguish between variants of greater and lesser impact. “Variant of concern” is an umbrella designation which indicates the possibility of adverse consequences but does not describe the probability or magnitude of those consequences. This may, in part, be a reflection of how challenging it is to quantify phenotypes like transmissibility, immune escape, and disease severity. Even with precise estimates, however, the leap from phenotype to population outcome can be complex.

The situation would be much more straightforward if variants existed in a vacuum, but they are part of a complex dynamical system. Variants may have selective advantages, but they are numerically disadvantaged by their late arrival; the extent to which a variant overcomes this disadvantage depends on the timing of its emergence as well as its phenotype. Vaccination complicates the picture even further: not only can it alter the balance between WT and variant, but it does so gradually, as vaccine rollout takes months. Finally, the timing of emergence differs between variants, and the speed of vaccine rollout varies between locations. Two variants with identical phenotypes can meet different fates simply by appearing at different times; the same variant may take off in one region and disappear from another.

The fact that population-level outcomes cannot be easily predicted from variant phenotypes does not mean that phenotypes are uninformative with respect to impact. Modeling can be used to examine outcomes for different variants across a range of scenarios, and comparing these outcomes – between variants and between scenarios – can inform our understanding of risk as well as the steps we take to reduce it. We describe here the results of a mathematical model which we use to simulate the emergence and spread of different variants in populations that are already mid-epidemic and controlling transmission through a combination of vaccination and nonpharmaceutical interventions (NPIs). We focus on three representative variants, which have enhanced transmissibility, partial immune escape, and a combination of both, respectively. We compare these variants in key outcomes, including the total number of infections and the number and percentage of infections averted by vaccination, highlighting the population-level impact of different variant phenotypes. We also explore how population outcomes are affected by the timing of vaccine rollout, illustrating the relative importance of vaccinating earlier or faster, and the sensitivity of different variants to these parameters. Finally, we explore the effects of decreasing vaccine efficacy, reducing vaccination coverage, or lifting NPIs once a certain percentage of the population is vaccinated, to compare the robustness of different variants to shortcomings in various control measures.

## Results

A full description of the model can be found in the Methods, but we include a short overview here to give context for the results. We describe the default model conditions and assumptions, but analyses varying many of these assumptions are included in the Results. The model is an extended Susceptible-Infected-Recovered (SIR) compartment model, which includes two strains – WT and variant – as well as vaccination. The WT is assumed to have a basic reproduction number of 2.5, which is reduced to 1.5 by nonpharmaceutical interventions (NPIs) that remain in place throughout the simulation. Both the variant and the vaccine are introduced at specified times midway through the epidemic, and the vaccine is distributed at a constant rate until 100% coverage is reached. The vaccine is assumed to be 95% effective against infection with WT, with efficacy against the variant proportional to cross-reactivity between the strains. Infection is assumed to confer sterilizing immunity against the infecting strain; protection against the other strain is proportional to the degree of cross-reactivity, which is assumed to be symmetric. The model is run for a simulated duration of three years (except where noted otherwise) and we assume no waning of immunity over this period. All simulations assume a population size of 100 million individuals.

The purpose of the model is to examine the impact of two key phenotypes – transmissibility and immune escape – on the population dynamics of emerging variants and the course of SARS-CoV-2 epidemics. In order to disentangle the effects of these traits, and to explore their interactions, we consider three hypothetical variants: Variant 1, with 60% increased transmissibility relative to WT; Variant 2, with 40% immune escape (60% cross-reactivity with WT); and Variant 3, with both of these phenotypes. We also model a null variant, designated Variant 0, which is effectively identical to WT and serves as a baseline for comparison with other variants. The characteristics of these hypothetical variants are laid out in Table 1.

**Table 1.** Hypothetical variants for the model, with transmissibility and cross-reactivity defined with respect to WT. Immune escape is defined as the complement of cross-reactivity (60% cross-reactivity = 40% immune escape).

|  |  | Transmissibility |  |
| --- | --- | --- | --- |
|  |  | Equal to WT | 60% increase over WT |
| Immune escape | 100% cross-reactivity with WT | Variant 0 | Variant 1 |
|  | 60% cross-reactivity with WT | Variant 2 | Variant 3 |

Some existing variants of concern may have higher transmissibility and/or less immune escape than reflected in the default parameter values. Since our findings suggest transmissibility has greater impact than immune escape, we deliberately use a conservative estimate of transmissibility and a generous estimate of immune escape to avoid reinforcing the result. Although 40% immune escape lies at the upper end of the range of estimates for existing variants, higher degrees of immune escape are theoretically possible, if perhaps difficult to evolve [39]. Analyses of variants with higher and lower degrees of immune escape are included in the Results.

Model behavior is analyzed in terms of numbers of infections, which when summed over the course of a simulation, we denote by epidemic size. We emphasize numbers of infections, rather than cases or individuals infected; the former denotes only those infections which are detected, while the latter ignores the fact that individuals may be infected more than once. We assume that epidemic size is roughly proportional to critical outcomes such as total hospitalizations and total deaths. We therefore use epidemic size as a key metric for comparison between variants and across different scenarios, although in some cases we distinguish between infections in susceptible and recovered/vaccinated individuals, as the latter are likely to have a milder clinical course.

### Ability to increase to high frequency is driven primarily by transmissibility, not immune escape

We start by examining the dynamics that result when the hypothetical variants are introduced into WT epidemics in the absence of vaccination. As expected, variant 0 (the null variant) behaves identically to WT, with growth and decline occurring at the same times and at the same rates (Fig 1A). In contrast, variants 1 and 3, which both have enhanced transmissibility, grow faster than WT and quickly rise to high frequency. Variant 2, which has a moderate degree of immune escape, remains at a low level, infecting far fewer people than WT over the course of the simulation (Fig 1B).

**Fig 1.**
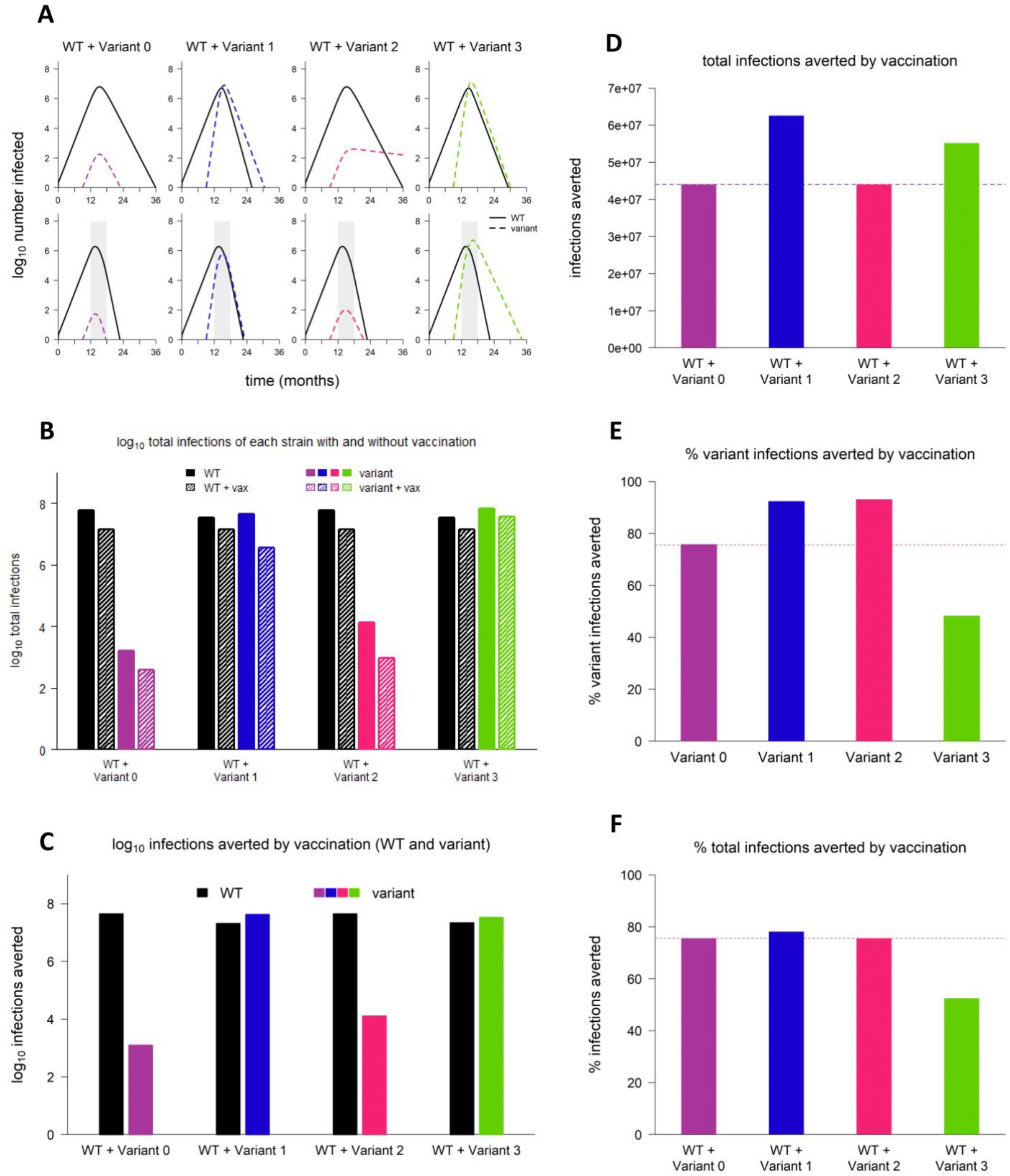
Sample dynamics of hypothetical variants. (A) Dynamics of WT and variant strains without vaccination (top row) and with vaccination (bottom row), shown on log scale. Solid/black lines, WT; dashed/colored lines, variants; gray shading, vaccine rollout. Subsequent panels reference the simulations depicted in (A). (B) Total infections with WT and variant strains with and without vaccination (log scale). Black bars, WT; colored bars, variants; solid bars, without vaccination; hatched bars, with vaccination. (C) WT and variant infections averted by vaccination (log scale). Black bars, WT; colored bars, variants. (D) Total infections (WT + variant) averted by vaccination (linear scale). Dashed line, total infections averted by vaccination in simulation with variant 0 (null variant). (E) Percentage of variant infections averted by vaccination (linear scale). Dashed line, percentage of varint infections averted in simulations with variant 0. (F) Percentage of all infections averted by vaccination (linear scale). Dashed line, percentage of infections averted in simulation with variant 0. In all simulations, variant is introduced at 9 months; in simulations with vaccination, vaccine rollout starts at 12 months and is spread over 6 months. Variant phenotypes are as follows: variant 0, identical to WT; variant 1, 60% greater transmissibility; variant 2, 40% immune escape; variant 3, 60% greater transmissibility and 40% immune escape.

When vaccination is added, variant 1 is rapidly controlled, as is variant 2, despite having partial immune escape. In contrast, although vaccination “flattens the curve” of variant 3 by reducing the growth rate, it is unable to completely control this variant, which has a combination of enhanced transmissibility and immune escape (Fig 1A).

### Variants do not necessarily undermine the population-level impact of vaccination

It is interesting to note that variants do not necessarily reduce the population-level impact of vaccination. When a variant increases the size of the epidemic, the number of infections that could be averted by vaccination also increases. In this instance, compared to simulations with the null variant (variant 0), vaccination averts a higher number of infections of variants 1, 2, and 3 (Fig 1C) and a higher number of infections overall (Fig 1D). For variants 1 and 2, vaccination also averts a higher percentage of variant infections (Fig 1E) and an equal or greater percentage of total infections (Fig 1F) compared to the null variant. Thus, although immune escape reduces vaccine efficacy for the individual, it does not follow that an emerging variant with partial immune escape will significantly reduce the ability of vaccination to limit case numbers in a whole population.

### With NPIs in place, moderate immune escape alone has little impact on final epidemic size and vaccination impact

We next vary the timing of vaccine rollout – changing both the time at which vaccination begins and the rate at which the rollout proceeds – to answer two key questions. First, how do variants with different phenotypes compare across a range of scenarios? And second, which aspect of vaccine rollout has a greater impact on population-level outcomes – vaccination start time or vaccination rate?

We find that, regardless of the timing of vaccine rollout, the total number of infections is nearly identical between variant 0 (the null variant) and variant 2, which has a moderate degree of immune escape (Fig 2A). The number and percentage of infections averted by vaccination are likewise similar between variant 2 and the null variant (Fig 2B-C). (Later, we show that these outcomes can change if control measures are significantly reduced or if the degree of immune escape is very high.) In contrast, variants 1 and 3, which have increased transmissibility, can significantly increase the total number of infections; the potential increase is particularly great for variant 3, which also has partial immune escape (Fig 2A). Paradoxically, in some scenarios, the number of infections averted by vaccination is greater for variants 1 and 3 than for the null variant (Fig 2B); this occurs because these variants substantially increase incidence, which raises the potential impact of preventive measures. However, the percentage of infections averted by vaccination is not significantly higher than the null variant in any of the scenarios examined, and is sometimes markedly lower, especially for variant 3 (Fig 2C).

**Fig 2.**
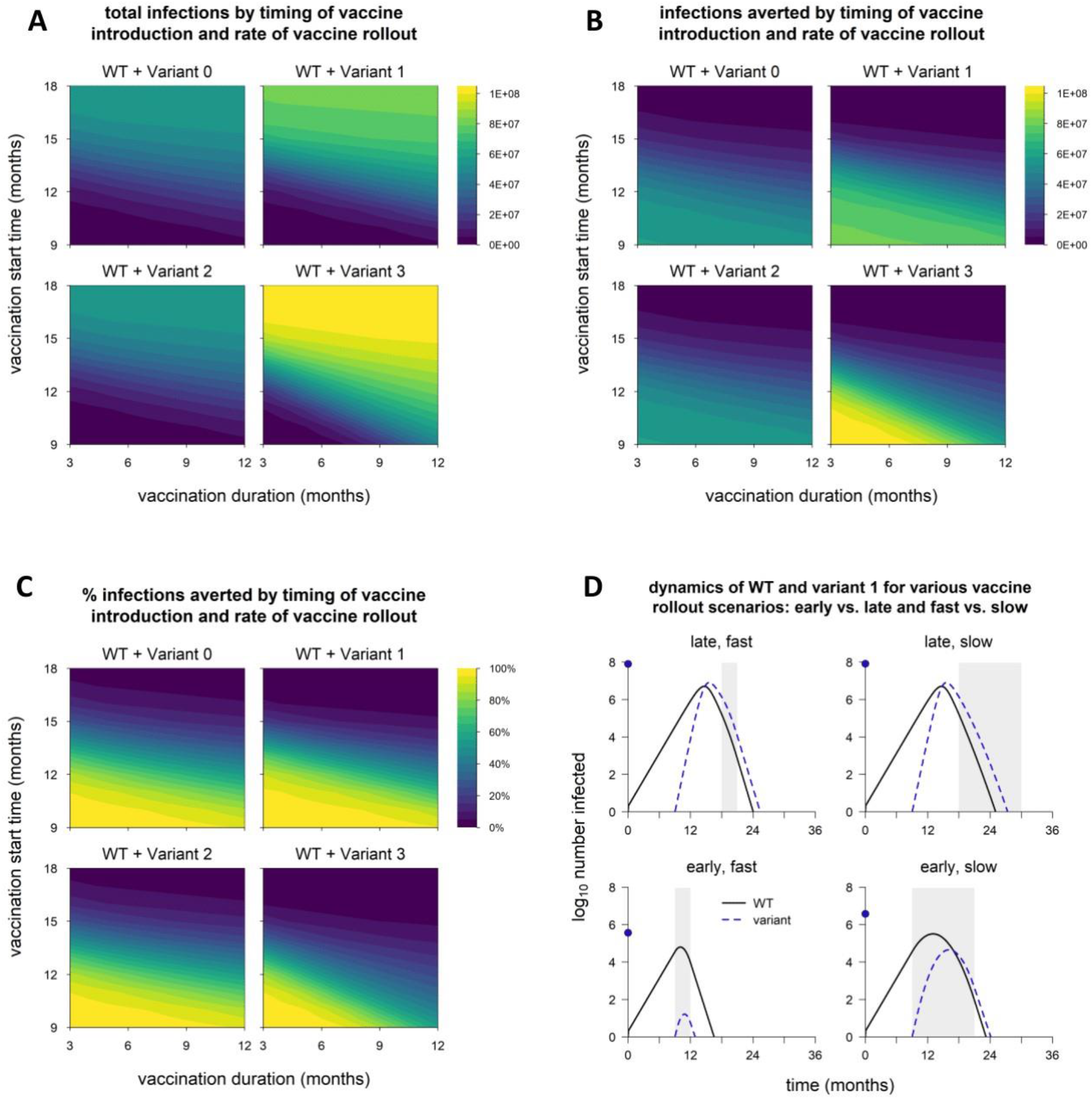
Epidemic size and vaccination impact vary with the time of vaccine introduction and rate of vaccine rollout. (A) Total infections (WT + variant) in simulations with each hypothetical variant, for varying rates of vaccination (vaccination duration, x-axis) and time of vaccine introduction (vaccination start time, y-axis); shaded contours represent total infections. (B) Number of infections (WT + variant) averted by vaccination; shading represents number of infections averted. (C) Percentage of infections (WT + variant) averted by vaccination; shading represents % infections averted. Variant introduced at 9 months in all simulations. (D) Dynamics of WT and variant 1 in simulations with the minimum and maximum values for vaccination duration (fast = 3 months, slow = 12 months) and start time (early = 9 months, late =18 months). In each panel, point on y-axis indicates the total number of infections over the entire simulation. Solid/black line, WT; blue/dashed line, variant; gray shading, vaccine rollout. Variant phenotypes are as follows: variant 0, identical to WT; variant 1, 60% greater transmissibility; variant 2, 40% immune escape; variant 3, 60% greater transmissibility and 40% immune escape.

### Population-level outcomes are more sensitive to vaccination start time than vaccination rate

For all variants, the total number of infections increases when the start of vaccination is delayed or the pace of vaccine rollout slows (Fig 2A). The reverse is true of vaccination impact: as vaccine rollout is delayed or slowed, the number and proportion of infections averted decreases (Fig 2B-C). However, all of these outcomes are more sensitive to vaccination start time than the rate of vaccine distribution; a one-month delay in starting vaccination changes outcomes more than a one-month increase in the duration of vaccine rollout. The sensitivity to vaccination start time is particularly pronounced for variants 1 and 3, which have increased transmissibility. Higher transmissibility increases the rate at which cases grow, which means the epidemic peaks earlier. A delay in starting vaccination can result in the peak of the epidemic being missed entirely, even if the pace of vaccine distribution is high (Fig 2D). However, if vaccination begins early, the effect on the final size of the epidemic can be considerable, even if the rollout is slow.

### Reinfections and breakthrough infections remain rare with moderate immune escape unless aided by transmissibility

In any scenario involving variants, especially variants with immune escape, it is important to consider how many infections occur in people with immunity from prior infection or vaccination, as a means of distinguishing between the potential to infect people with immunity and the actual occurrence of such infections. In addition, infections in those with prior immunity are likely to be less severe than infections in naïve individuals, and distinguishing between infections in naive and recovered/vaccinated hosts may better reflect the severity of an epidemic. We therefore separated infections into those in people with prior infection and/or vaccination and those in naive individuals (no history of infection or vaccination). Infections in naïve hosts exhibit the same patterns as the total infections (Fig 2A, Fig 3A), but infections in recovered and vaccinated individuals exhibit different behavior.

**Fig 3.**
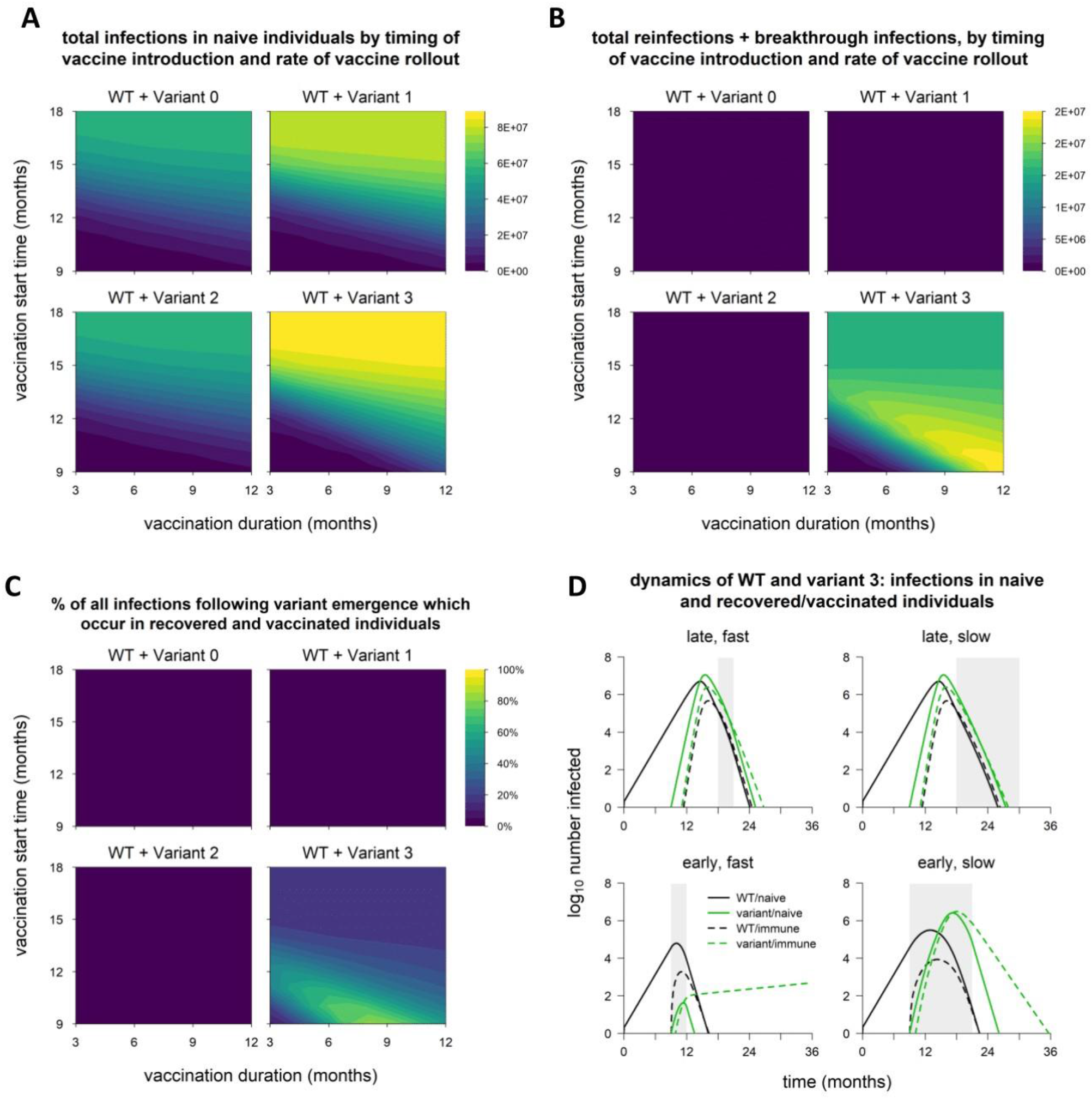
Breakdown of infections by immune status (naïve vs. recovered/vaccinated). (A) Total infections in naïve individuals, in simulations with each variant, for varying rates of vaccination (x-axis) and time of vaccine introduction (y-axis); shaded contours represent total infections. (B) Total infections in recovered and vaccinated individuals (reinfections and breakthrough infections); shaded contours represent total infections. (C) Percentage of all infections occurring in recovered/vaccinated individuals (reinfections/breakthrough infections), starting from the beginning of vaccine rollout; shading represents percentage of infections occurring in recovered/vaccinated individuals. Variant introduced at 9 months in all simulations. (D) Dynamics of WT and variant 3, broken down by host immune status (naïve vs. recovered/vaccinated, in simulations with the minimum and maximum values for vaccination duration (fast = 3 months, slow = 12 months) and start time (early = 9 months, late =18 months). Black lines, WT; green lines, variant; solid lines, infections in naïve individuals; dashed lines, infections in recovered/vaccinated individuals; gray shading, vaccine rollout. Variant phenotypes are as follows: variant 0, identical to WT; variant 1, 60% greater transmissibility; variant 2, 40% immune escape; variant 3, 60% greater transmissibility and 40% immune escape.

Reinfections and breakthrough infections (which we define as active infections in previously infected and vaccinated individuals, respectively) are negligible in simulations with variants 1 and 2, which suggests that neither enhanced transmissibility nor a moderate degree of immune escape will necessarily lead to significant numbers of reinfections/breakthrough infections (Fig 3B-C). However, a combination of partial immune escape and increased transmissibility can produce significant numbers of reinfections and breakthrough infections, even accounting for a majority of infections following emergence of the variant (Fig 3C). In this case, the number of reinfections/breakthrough infections is maximized when vaccine rollout starts early but distribution is slow. Counterintuitively, this reinforces the benefits of beginning vaccine rollout early rather than maximizing the rate of distribution. If either speed or promptness is to be sacrificed, reducing the speed of the rollout will cause fewer excess infections, and additional infections that do arise will be more likely to occur in previously infected/vaccinated individuals, who tend to have milder illness (Fig 3D).

### Weakening control measures leads to substantial excess infections from variants with enhanced transmissibility

We now explore how the course of the epidemic is affected by weakening control measures in various ways. We first evaluate the effects of lifting nonpharmaceutical interventions (NPIs) once vaccination coverage reaches 50%. For variants 1 and 2, the number of excess infections from lifting NPIs is fairly small (Fig 4A, Fig 5B,C). For variant 3, however, lifting NPIs can lead to a significant increase in the final size of the epidemic, especially when vaccine rollout is early and/or rapid; efficient vaccine rollout prevents a larger number of cases, which increases the potential rebound following relaxation of NPIs (Fig S1).

**Fig 4.**
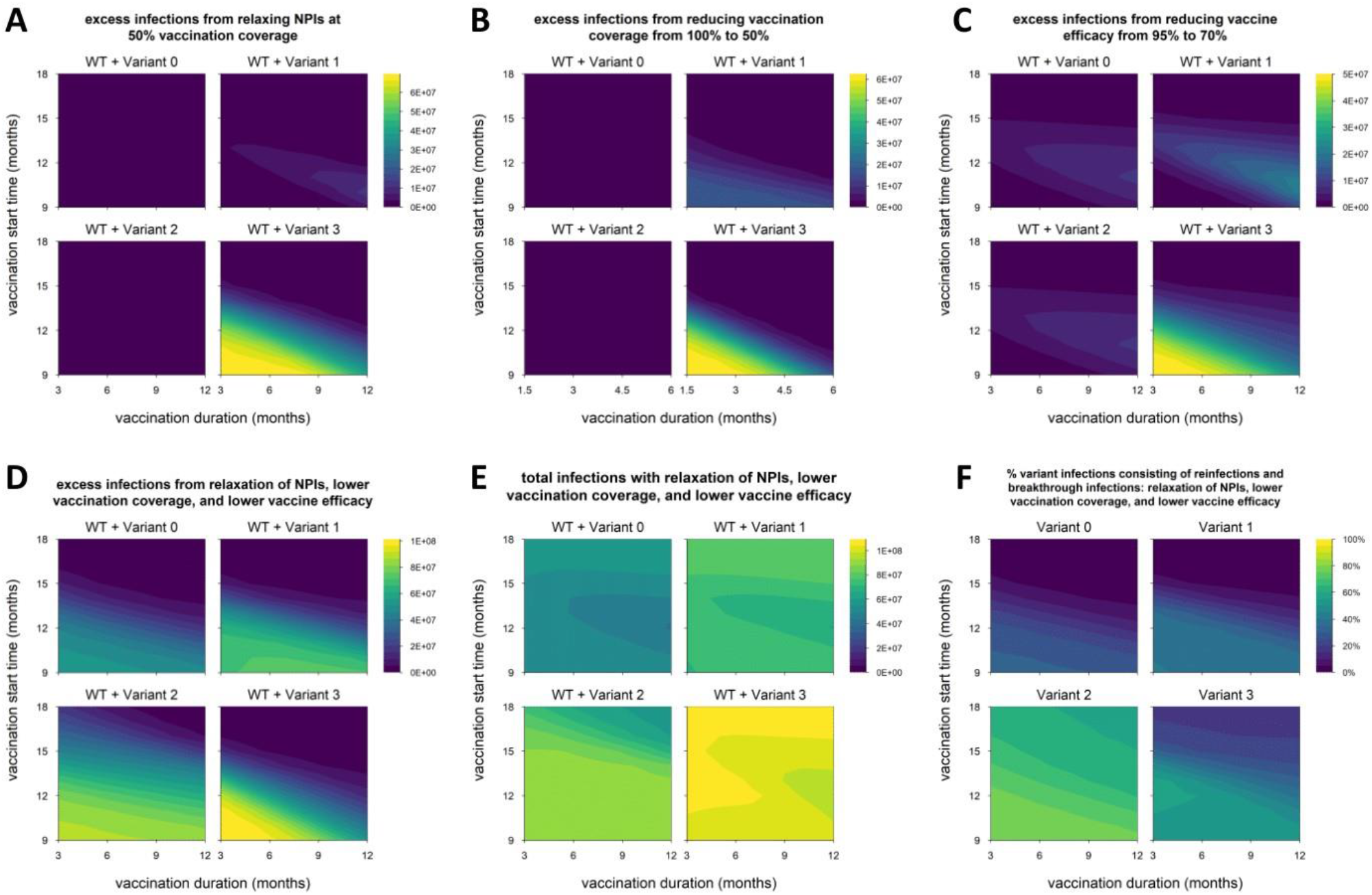
Impact of reduced control measures on epidemic size. (A) Excess infections from lifting nonpharmaceutical interventions (NPIs) when vaccination coverage reaches 50% (default condition is NPIs continued indefinitely); shading represents excess infections compared to default conditions/parameters. (B) Excess infections from reducing vaccination coverage from 100% to 50%. (C) Excess infections from reducing baseline vaccine efficacy (against WT) from 95% to 70%. (D) Excess infections from the combination of conditions A through C (lifting NPIs, reduced coverage and reduced efficacy). (E) Total infections (WT + variant) under the combined conditions of panels A through C; shading represent total infections. (F) Percentage of variant infections composed of reinfections and breakthrough infections under the combined conditions of panels A through C; shading represent percentage of variant infections occurring in recovered/vaccinated individuals. Variant introduced at 9 months in all simulations. Variant phenotypes are as follows: variant 0, identical to WT; variant 1, 60% greater transmissibility; variant 2, 40% immune escape; variant 3, 60% greater transmissibility and 40% immune escape.

**Fig 5.**
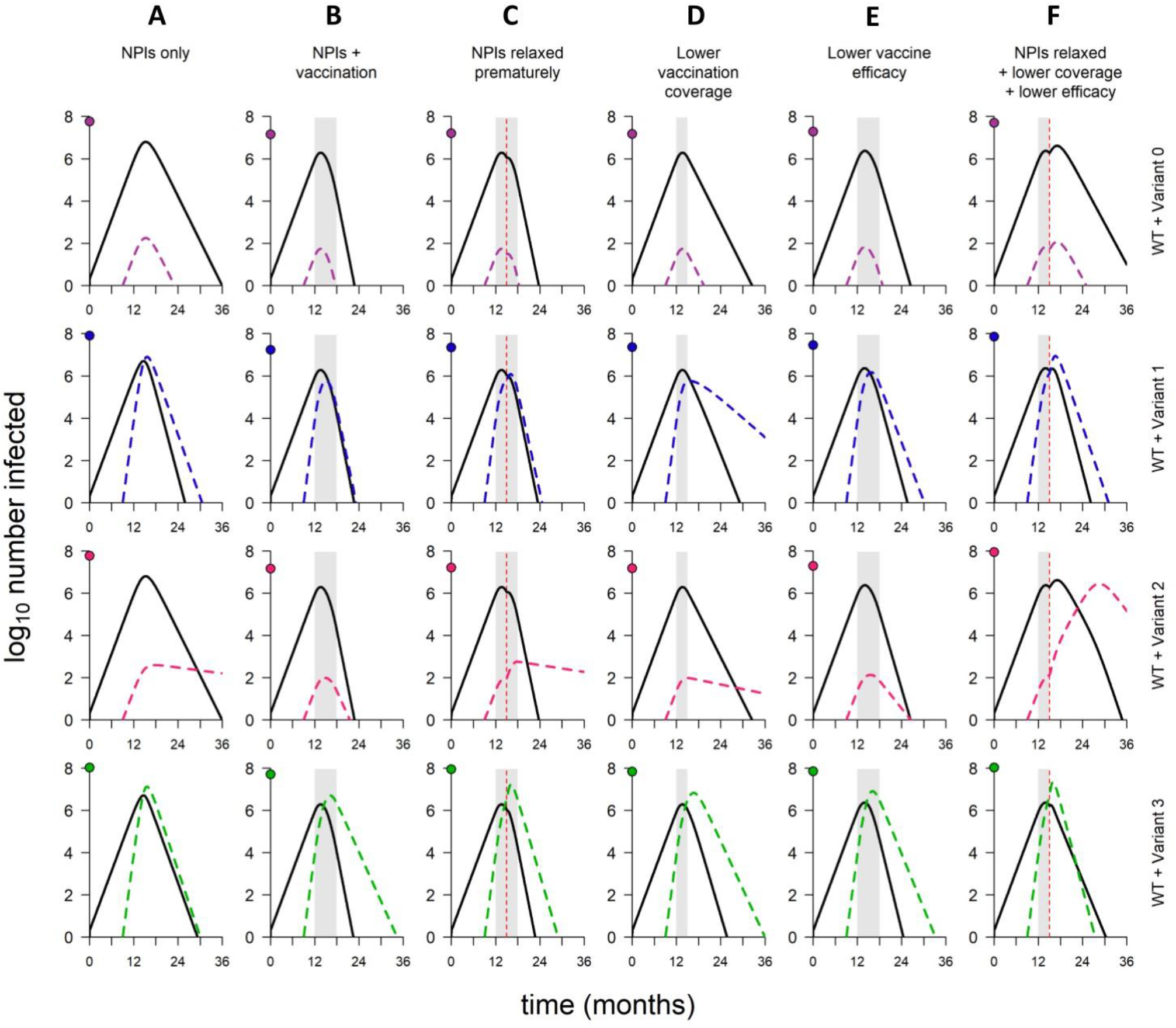
Dynamics of WT and variants in simulations with varying combinations of control measures. (A) No vaccination but NPIs in place throughout. (B) Default model conditions (NPIs in place throughout, 100% vaccination coverage, 95% vaccine efficacy against WT). (C) NPIs lifted when vaccination coverage reaches 50% (other model conditions as in B). (D) 50% vaccination coverage (other model conditions as in B). (E) 70% vaccine efficacy (other model conditions as in B). (F) Combination of conditions C through E (NPIs relaxed, 50% vaccination coverage, and 70% vaccine efficacy). In each panel, point on y-axis indicates the total number of infections over the entire simulation. Variant introduced at 9 months in all simulations; in simulations with vaccination, rollout begins at 12 months and lasts 6 months if final coverage is 100%, 3 months if final coverage is 50%. Solid/black lines, WT; colored/dashed lines, variants; gray shading, vaccine rollout; dashed vertical line, 50% vaccination coverage. Variant phenotypes are as follows: variant 0, identical to WT; variant 1, 60% greater transmissibility; variant 2, 40% immune escape; variant 3, 60% greater transmissibility and 40% immune escape.

We next consider the impact of reducing vaccination coverage from 100% to 50%. Surprisingly, this does not provide a significant advantage to variant 2 (partial immune escape) and does not lead to an appreciable increase in the number of infections (Fig 4B, Fig 5B,D). However, reducing coverage can notably increase the number of infections in simulations with variant 1 or 3, since the critical vaccination threshold for these more transmissible variants is higher. For both of these variants, the increase in infections is particularly pronounced when vaccination is early and/or rapid (Fig S2), for the same reasons described above.

Finally, we examine the consequences of lowering baseline vaccine efficacy (against WT) from 95% to 70%. Here, too, the impact on epidemic size is minimal in simulations with variant 2 (Fig 4C, Fig 5B,E), suggesting that variants with moderate degrees of immune escape may be suppressed even with suboptimal levels of vaccine coverage or efficacy (Fig S2, S3). However, variant 3, which has enhanced transmissibility as well as partial immune escape, can cause a substantial increase in the number of infections when vaccine efficacy is reduced, especially with early/rapid rollout. The cost of lower efficacy can also be considerable with variant 1; however, in this case, the number of excess infections is greatest when vaccine rollout is slow. In contrast with variant 3, high coverage with a less effective vaccine is sufficient to control variant 1; therefore, the cost of lower efficacy is determined by what happens during vaccine rollout, rather than after. Slower rollout provides a longer period of time during which infections continue to increase, resulting in a larger number of excess infections (Fig S3).

### Sufficiently weak control measures can lead to a second wave of infections with immune escape variants

We also explore the impact of combining the three changes described above – reducing vaccination coverage to 50%, lowering vaccine efficacy to 70%, and lifting NPIs when coverage reaches 50%. Unsurprisingly, this leads to many additional infections (Fig 4D). However, in contrast with the scenarios considered above, surprisingly large numbers of excess infections are observed in simulations with variant 2, with the totals frequently exceeding those of simulations with the more transmissible variant 1 (Fig 4E). The dramatic increase in infections observed in simulations with variant 2 is driven by a “second wave” of the variant, which is still able to spread even as the WT declines (Fig 5F). Although the second wave substantially increases the total number of infections, the majority of variant 2 infections are reinfections and breakthrough infections, which are likely to be milder than infections in naïve individuals (Fig 4F).

### Higher levels of immune escape increase the propensity for a second wave of variant infections

We next examine the impact of lower and higher levels of immune escape; the former probably exists among existing variants of concern, while the latter is, at present, only a theoretical possibility. For a lower degree of immune escape, variants 2 and 3 are assumed to have 20% escape (80% cross-reactivity with WT); for a higher degree, we assume 80% escape (20% cross-reactivity) for these variants. In both cases, we assume variants 1 and 3 are 60% more transmissible than WT (as in all other simulations). In general, we find that a lower degree of immune escape does not qualitatively change our findings (Fig S4-S7), except that multiple reductions in control measures (reduced vaccine coverage, reduced vaccine efficacy, and lifting NPIs midway through vaccine rollout) does not lead to a second wave with variant 2, resulting in fewer reinfections/breakthrough infections and fewer infections in total.

In contrast, a very high level of immune escape significantly enhances the spread of variants 2 and 3 and limits the ability of vaccination to control these variants (Fig S8-S11). When variant 2 has a very high level of immune escape, the total epidemic size is noticeably higher than with variant 0 (the null variant), although the epidemic size is often still greater with the more transmissible variant 1 (Fig S8A). In simulations with variant 2, the increase in epidemic size is driven by a second wave of variant infections following the initial wave of WT infections (Fig S11); in simulations with variant 3, rather than a second wave, the waves of WT and variant infections peak almost simultaneously. Paradoxically, in simulations with variant 2, vaccination still averts a larger number of infections, and sometimes a greater percentage of infections, than in simulations with variant 0, even though vaccine efficacy against the variant is very low (Fig S8B-C). This occurs because vaccination averts a similar number of WT infections as well as a small proportion of the greatly increased number of variant infections. However, in simulations with variant 3, vaccination is unable to control the variant’s rapid spread and accordingly averts a low number and small percentage of infections.

With a high level of immune escape, variant 2 is not controlled even with optimal interventions, but NPIs and vaccination have enough impact that lifting NPIs or reducing vaccine coverage or efficacy leads to a significant increase in the total number of infections, although many of the variant infections are reinfections and breakthrough infections (Fig S10-S11). Whereas interventions help blunt the impact of variant 2, their effectiveness against variant 3 is minimal, such that total infections increase only slightly when control measures are reduced. NPIs and vaccination can therefore have an appreciable impact in the presence of a variant with a high degree of immune escape, but if combined with enhanced transmissibility, more stringent measures might be needed to limit transmission.

### Qualitative findings are largely robust to changing key model assumptions

Finally, we re-examine all of the results discussed above after varying two key model assumptions. First, we change the way non-pharmaceutical interventions are implemented in the model. By default, we assume NPIs are fixed to achieve a constant 40% reduction in the reproduction number. This simplification generates dynamics that are easy to interpret and analyze; however, the dynamics of SARS-CoV-2 in most settings have been characterized by multiple waves. These waves are caused, in large part, by switching between lax and stringent control measures, as dictated by rising and falling case numbers. We developed an alternative version of the model, in which the virus is kept in check by rolling lockdowns that come into effect whenever prevalence exceeds a set threshold (Fig S12). The findings with this alternative model are largely similar to those of the default model (Fig S13-S15), except that variant 1 does not perform as well, especially compared to variant 3. In simulations with variant 1, the total number of infections is more similar to simulations with the null variant (Fig S13A), and the number of excess infections resulting from reduction of control measures is minimal except when multiple aspects of control measures are weakened simultaneously (Fig S15A-D). This is because sufficiently strong control measures (i.e. lockdowns) can keep even highly transmissible variants in check, especially when complemented by vaccination and cross-immunity. The degree to which this occurs in practice will depend on the strength of the control measures, the responsiveness of control measures to case numbers (e.g. thresholds and lags for implementation), and the transmissibility of the variant(s) present.

Second, we changed a core assumption about the way immunity limits infection. The default model assumes that immunity reduces the probability of infection by decreasing the rate of movement from uninfected to infected states (so-called “leaky” immunity). An alternative construction assumes that a given individual either does or does not develop immunity to a given strain following infection or vaccination (termed “all-or-nothing” immunity). Because these can theoretically give rise to different outcomes, we re-ran all simulations using an alternative version of the model in which immunity behaves in an all-or-nothing manner. The results are qualitatively nearly indistinguishable, but the numbers of infections are generally lower with all-or-nothing immunity (Figs S16-S18), since some fraction of all recovered and vaccinated individuals are completely refractory to infection.

## Discussion

In this work, we use a mathematical model to simulate the spread of SARS-CoV-2 variants with different phenotypes and explore how nonpharmaceutical interventions and vaccination affect the resulting dynamics. We show that a moderate degree of immune escape, on its own, is a relatively minor concern: variants with this phenotype may fail to overtake the WT strain, and if this does occur, infections will disproportionately occur in previously infected and vaccinated individuals, who are less likely to suffer severe disease. In contrast, variants with enhanced transmissibility are able to invade under most circumstances and may considerably increase the size of the epidemic (i.e. the total number of infections). Furthermore, whereas partial immune escape is frequently inconsequential on its own, when combined with enhanced transmissibility, immune escape limits the ability of vaccination to bring an epidemic under control because the variant rapidly becomes the dominant strain in the population. Thus, a moderate level immune escape can lead to severe consequences, but is unlikely to do so unless paired with enhanced transmissibility.

In this context, it is important to note that transmissibility should be evaluated with respect to the dominant strain in the population, rather than a particular “original” strain. For instance, in our model, variant 3 has increased transmissibility as well as partial immune escape, and this variant easily invades when WT is the dominant strain. However, this variant would generally fail to invade against variant 1, since these variants are equally transmissible. Thus, the acquisition of partial immune escape on a highly transmissible background may be inconsequential if the highly transmissible lineage is already the dominant strain.

These findings advance our understanding of the dangers posed by different variants. Currently, both the World Health Organization and the US Centers for Disease Control and Prevention designate certain strains as “variants of concern” using similar criteria, which include evidence of immune escape, vaccine escape, and/or enhanced transmissibility. In general, variants are not assigned threat levels or differentiated status, although the US CDC would recognize variants with clear evidence of significant vaccine escape as “variants of high consequence.” Our findings suggest that increased transmissibly may be considered a red flag which signals a strong tendency to invade and the potential to considerably increase numbers of infections. Partial immune escape, absent enhanced transmissibility, is likely to have mild consequences in comparison. Even if a considerable degree of immune escape were to emerge on a highly transmissible background, such as Alpha or Delta, the impact may be negligible if the predecessor is already the most abundant strain in the population.

Our findings also show that variants reinforce the importance of vaccination without necessarily diminishing vaccination’s impact. Because variants - especially those with enhanced transmissibility - increase the total size of the epidemic, vaccination often prevents a larger number of infections and/or a greater percentage of total infections when variants are present. Even in the case of high levels of immune escape, vaccination may substantially improve outcomes compared to the alternative of no vaccination. However, variants also increase the consequences of imperfect control measures, for several reasons. If vaccination prevents more infections in the presence of variants, then the impact of reduced vaccination is, conversely, greater. In addition, higher levels of vaccine coverage and vaccine efficacy are required to control variants with enhanced transmissibility and/or partial immune escape, which means a modest reduction in coverage or efficacy may result in failure to control the epidemic. Finally, we found that, for all strains, epidemic size is more sensitive to the time at which vaccine rollout is started than the rate at which rollout proceeds. However, this effect was more pronounced for variants with enhanced transmissibility; a higher reproduction number accelerates the entire epidemic, which necessitates earlier intervention to limit transmission.

These results provide a theoretical basis to understand the behavior of existing variants, predict the behavior of future variants, and design appropriate strategies to mitigate the impact of variants of concern in populations across the world. Our work may explain why highly transmissible variants Alpha and Delta rapidly swept to near-fixation while Beta, which probably has the highest degree of immune escape among existing variants, largely failed to do so. In addition, our findings suggest that evolution of partial immune escape on strain backgrounds that are already common (e.g. Delta) may be difficult, even if the relevant mutations occur frequently. Lastly, our work underscores the importance of beginning vaccine rollout on a global scale, as soon as possible, to mitigate the impact of highly transmissible variants of concern.

### Limitations of the study

An obvious limitation of our model is that it does not allow for circulation of multiple variants in the same population. In reality, variants compete with one another as well as the WT strain, and this may lead to complex behavior that is difficult to predict from pairwise interactions. It is particularly challenging to anticipate the dynamics of a system with multiple variants that each have some degree of enhanced transmissibility and immune escape, as the relative fitness of these strains may flip as the level of immunity in the population increases. Additional modeling is therefore required to understand the threats posed by different variants circulating simultaneously in a single population.

Our model is also strictly focused on what happens to a variant following emergence; the phenomenon of emergence itself, which is inherently stochastic, is not considered here. The generation and persistence of novel lineages is related to the size of the infected population, as well as the selective pressures acting on the viral population. The emergence of new variants is therefore affected by control measures, such as nonpharmaceutical interventions and vaccination, that limit the prevalence of infection and alter the strength of selection on different traits [40]. This will be an important dimension to consider in future work, as the optimal strategies to minimize transmission of a particular variant may or may not be aligned with the best strategies to limit emergence.

We also do not explicitly consider the clinical side of the equation. In addition to differences in severity between naïve and recovered/vaccinated individuals, we do not take into account the possibility of variants with increased propensity to cause severe disease, as has been reported for Delta [41, 42]. Differences in severity affect estimates of cumulative morbidity and mortality, but also impact the burden on healthcare systems. Large numbers of severe cases, which may be attributable to more virulent strains or simply uncontrolled transmission, can stretch clinical resources thin, with negative consequences for patient outcomes. Further work is needed to combine studies of clinical outcomes with epidemiological modeling, including utilization of healthcare resources, to estimate potential morbidity and mortality for specific variants of concern.

Finally, there would be value in extending the model used here, or subsequent models, to multiple populations. The issue of how to optimize vaccine rollout, for example, may be extended to consider strategies for vaccine allocation across different populations. This is particularly relevant for models that include stochastic emergence of novel variants, as emergence of a variant of concern in one population will eventually threaten all populations. Such work would have relevance for the ongoing distribution of COVID-19 vaccines as well as strategic preparation for future pandemics.

## Methods

### Model overview

We use an ordinary differential equation (ODE) compartment model to simulate the dynamics of WT and variant strains of SARS-CoV-2 in the context of nonpharmaceutical interventions (NPIs) and vaccine rollout. The model is an extended version of the standard SIR (susceptible-infected-recovered) model framework with two key changes. First, the model tracks two viral strains, which are denoted strain 1 (WT) and strain 2 (variant). Second, the model allows for strain-specific immunity to be acquired via infection and vaccination. In addition, the model is implemented with a wrapper that enables various events, such as variant emergence, vaccine rollout, or intensifying/relaxing control measures, to occur at pre-specified times and/or in response to the state of the system.

### Infection states

In this compartment model, individuals are classified according to their current infection status as well as their infection and vaccination history. Four types of compartments exist: *S* (susceptible), *I* (infected), *R* (recovered), and *V* (vaccinated). Subscripts further distinguish between infection states and infection histories within the *I, R*, and *V* classes (Table 2).

**Table 2.** State variables and definitions.

| State variable | Definition |
| --- | --- |
| $S$ | Susceptible |
| $I_{iS}$ | Infected with strain $i$, previously susceptible |
| $I_{iS(V)}$ | Infected with strain $i$, previously susceptible, vaccinated during current infection |
| $R_i$ | Recovered from strain $i$ |
| $I_{iR}$ | Infected with strain $i$, previously recovered (from strain $j$) |
| $I_{iR(V)}$ | Infected with strain $i$, previously recovered (from strain $j$), vaccinated during current infection |
| $R_{ij}$ | Recovered from both strain $i$ and strain $j$ |
| $V$ | Vaccinated (never infected) |
| $I_{iV}$ | Infected with strain $i$, previously vaccinated |
| $V_i$ | Vaccinated and recovered from strain $i$ |
| $I_{iV(j)}$ | Infected with strain $i$, previously vaccinated, previously recovered from strain $j$ |
| $V_{ij}$ | Vaccinated and recovered from both strain $i$ and strain $j$ |

Movement between compartments is the result of three processes: infection, recovery, and vaccination. Infection is a mass-action process resulting from contact between infected individuals and susceptible individuals (meaning all who are not completely refractory to infection). All individuals infected with a given strain are assumed to be equally infectious, with transmission rate *β* for WT and (1 + *λ*)*β* for the variant (see *Variant phenotypes*). All infected individuals also have the same average duration of infectiousness, which is the inverse of the recovery rate, *γ* (Table 3). Individuals may have varying susceptibility to each strain, depending on prior infection and/or vaccination. Upon recovery, individuals develop sterilizing immunity against the infecting strain and partial protection against the non-infecting strain; the degree of cross-protection is given by *ϕ*.

**Table 3.** Model parameters, definitions, and default values.

| Parameter | Definition | Default value |
| --- | --- | --- |
| $N$ | Population size | $10^8$ |
| $R_0$ | Reproduction number (WT) | 2.5 |
| $\gamma$ | Recovery rate | $1/14 \text{ days}^{-1}$ |
| $\beta$ | Transmission rate | $\frac{R_0 \gamma}{N}$ |
| $\phi$ | Cross-reactivity between WT and variant | See Tables 4-5 |
| $\omega$ | Maximum vaccine efficacy | 0.95 |
| $\alpha_1$ | Antigenic mismatch between vaccine and WT | 0 |
| $\alpha_2$ | Antigenic mismatch between vaccine and variant | $1 - \phi$ |
| $\kappa_I$ | Eligibility of infected individuals for vaccination; 1 = eligible, 0 = ineligible | 1 |
| $\kappa_R$ | Eligibility of recovered individuals for vaccination; 1 = eligible, 0 = ineligible | 1 |
| $\lambda$ | Increase in transmissibility of variant relative to WT | See Tables 4-5 |
| $\tau_2$ | Time of variant introduction | 9 months |
| $\tau_{vax}$ | Start of vaccine rollout | Varies |
| $v$ | Daily per-capita vaccination rate | Varies |
| $c$ | Final vaccination coverage | 100% |
| $r$ | Reduction in transmission due to nonpharmaceutical interventions | 0.4 |

Once vaccine rollout is initiated (see *Simulation Overview*), a fixed number of individuals are vaccinated per day, but these individuals are drawn only from eligible compartments. By default, all unvaccinated individuals are eligible. The parameters *κ*_*I*_ and *κ*_*R*_ control the vaccine eligibility of infected and recovered individuals, respectively (Table 3). Vaccination takes place in a single dose and is assumed to take effect immediately, unless the recipient is currently infected, in which case the vaccine takes effect upon recovery. The vaccine is assumed to have maximum efficacy *ω* against a perfect antigenic match. The degree of antigenic mismatch between the vaccine and strain *i* is given by *α*_*i*_.

Both infection-induced and vaccine-induced immunity are assumed to be durable, lasting longer than the duration of the simulations; waning of immunity is not included in the model.

### Variant phenotypes

Variants are characterized in terms of two phenotypes: transmissibility and immune escape. The increase in transmissibility relative to WT is denoted by *λ*, giving a transmission rate of (1 + *λ*)*β*. Immune escape is the complement of cross-reactivity between WT and variant (1 – *ϕ*). The default phenotypes of the hypothetical variants modeled are given in Table 4.

**Table 4.** Hypothetical variant phenotypes

|  |  | Increased transmissibility |  |
| --- | --- | --- | --- |
| | | $\lambda = 0$ | $\lambda = 0.6$ |
| Partial immune escape | $\phi = 1$ | Variant 0 (control) | Variant 1 (more transmissible) |
| | $\phi = 0.6$ | Variant 2 (partial escape) | Variant 3 (more transmissible + partial escape) |

### Simulation overview

Each simulation begins with a single WT infection introduced into a susceptible population of size *N*. The variant is introduced at time *τ*_2_, and rollout of the vaccine begins at time *τ*_*vax*_. Vaccination proceeds at a constant rate, with a fixed proportion *v* of the population vaccinated per day until the maximum coverage *c* is reached (the duration of vaccine rollout is thus 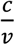). The total length of each simulation is three years, except for simulations with an increased level of immune escape, which were run for a simulated duration of six years.

### Nonpharmaceutical interventions

NPIs are assumed to be in place throughout the simulation, except in some scenarios where NPIs are lifted once vaccination coverage reaches 50%. These control measures are assumed to reduce transmission of both strains by a factor *r*.

### Simulations

Simulations were run with each variant alone and in combination with WT; the latter category of simulations were run with and without vaccination (Table 5). For each set of simulations, the start of vaccine rollout (*τ*_*vax*_) and the duration of vaccine rollout 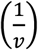 were varied in one-month increments over the ranges given in Table 5.

**Table 5.** Simulations under default model configuration (parameter values as listed in Table 3). Variants 0, 1, etc. are abbreviated V0, V1, etc.

| Set | Strain(s) | Start of vaccine rollout ($\tau_{vax}$) | Duration of vaccine rollout ($\frac{1}{v}$) | Variant transmission advantage ($\lambda$) | Cross-reactivity ($\phi$) |
| --- | --- | --- | --- | --- | --- |
| 1 | V0 | NA | NA | 0 | 0 |
| 2 | V1 | NA | NA | 0.6 | 0 |
| 3 | V2 | NA | NA | 0 | 0.6 |
| 4 | V3 | NA | NA | 0.6 | 0.6 |
| 5 | W + V0 | NA | NA | 0 | 0 |
| 6 | W + V1 | NA | NA | 0.6 | 0 |
| 7 | W + V2 | NA | NA | 0 | 0.6 |
| 8 | W + V3 | NA | NA | 0.6 | 0.6 |
| 9 | W + V0 | 9-18 mos | 3-12 mos | 0 | 0 |
| 10 | W + V1 | 9-18 mos | 3-12 mos | 0.6 | 0 |
| 11 | W + V2 | 9-18 mos | 3-12 mos | 0 | 0.6 |
| 12 | W + V3 | 9-18 mos | 3-12 mos | 0.6 | 0.6 |

### Model equations

We use the shorthand *p*(*t*) for the per capita rate of vaccination among the eligible population. Once vaccination is initiated, a fraction *v* of the population is vaccinated daily (*Nv* in absolute numbers), but because the eligible population fluctuates in size, it is necessary to calculate the rate at which individuals in eligible model compartments are vaccinated. We define *E*(*t*) as the eligible population at time *t*, and calculate *E*(*t*) as follows:

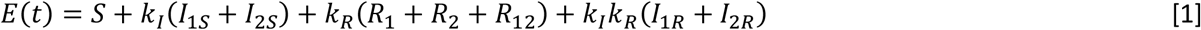

We then define *p*(*t*) as follows:

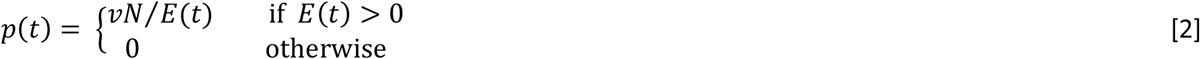

In addition, because there are six infected compartments for each strain, we simplify the equations by defining new state variables representing the total number of infected individuals for each strain:

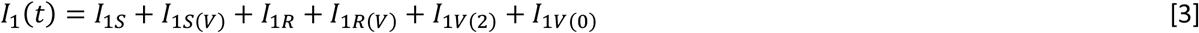

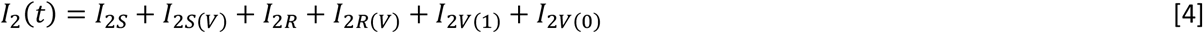

The rates of movement between compartments are shown alongside the model diagram in Fig 6. The model equations are given by Equations 5-24.

**Fig 6.**
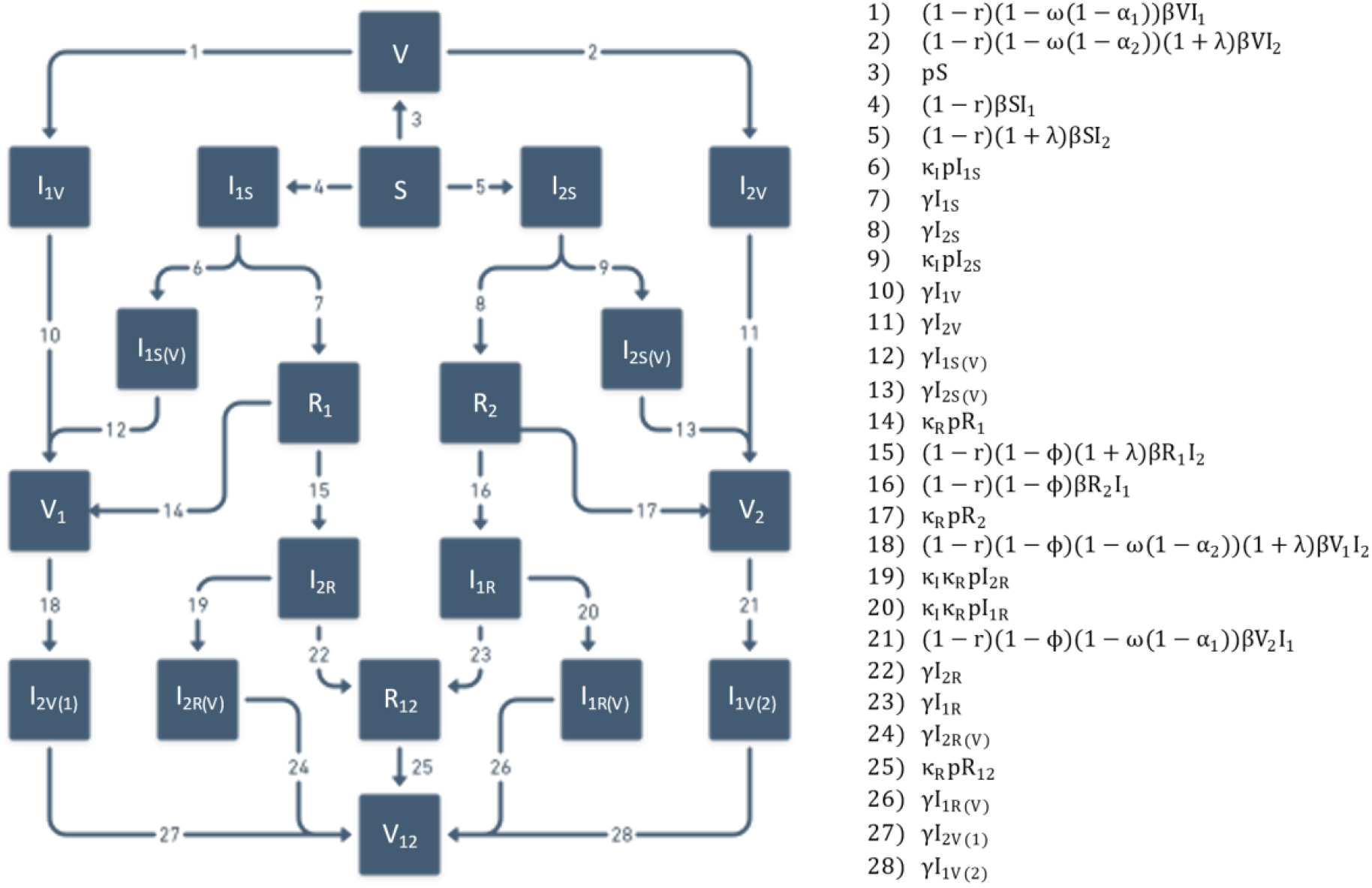
Model diagram and rates of movement between compartments, corresponding to labeled arrows.

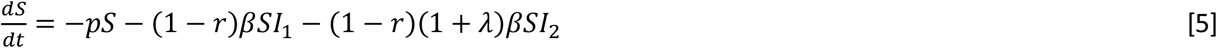

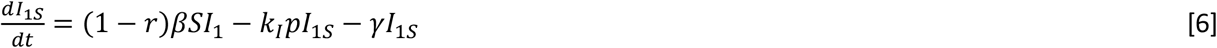

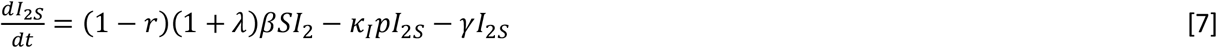

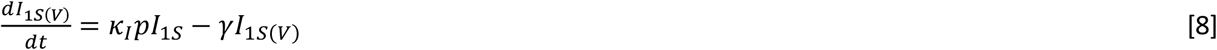

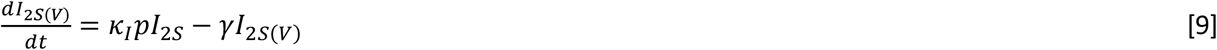

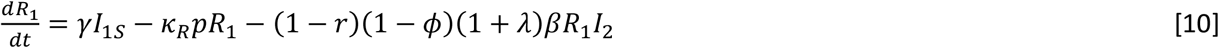

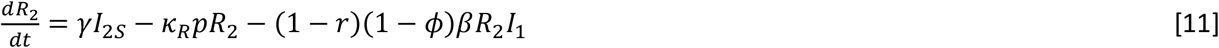

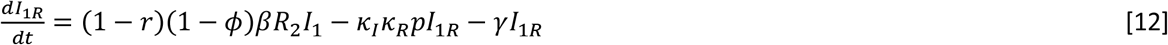

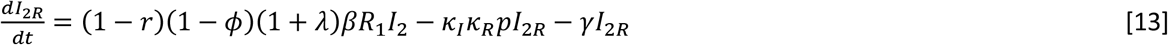

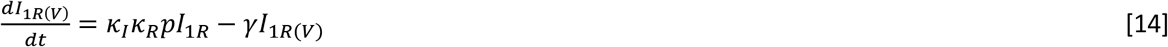

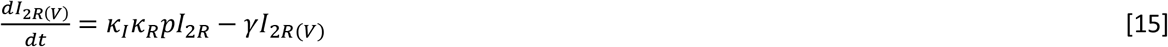

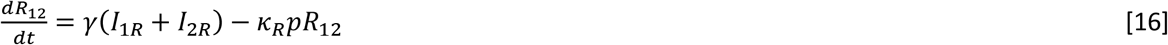

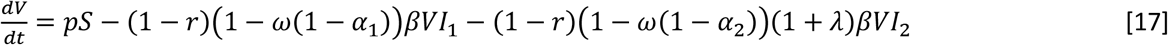

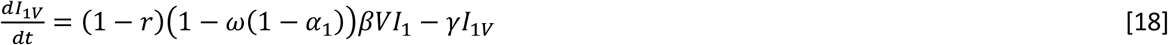

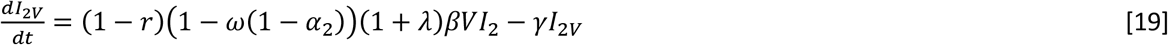

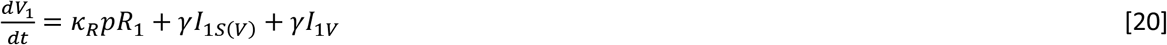

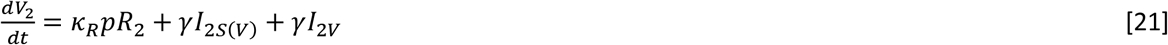

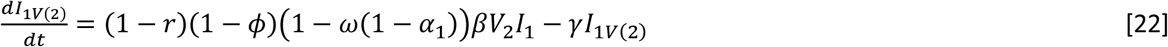

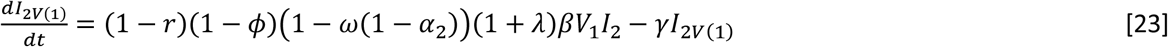

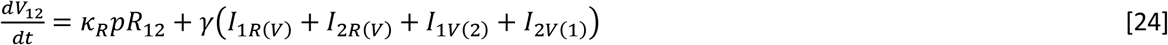

### Model outcomes

The main outcome compared across simulations is the cumulative number of infections, which is modeled as the sum of flows into all infected compartments. Similar quantities are defined for infections in naïve individuals (previously belonging to compartment *S*) and in recovered/immune individuals (coming from *R* and *V* compartments). The relative frequency of reinfections and breakthrough infections is obtained by dividing the number of infections in recovered/immune individuals by the total number of infections in each simulation; this is reported only for the time period following variant emergence.

Vaccine impact is obtained by comparing numbers of infections between simulations with vaccination (sets 9-12 in Table 5) to otherwise identical simulations without vaccination (sets 5-8). If we denote the total number of infections with and without vaccination by *Y*_*vax*_ and *Y*_*novax*_, respectively, vaccine impact can be measured in terms of the number of infections averted (*Y*_*novax*_ – *Y*_*vax*_) or the percentage of infections averted (1 – *Y*_*vax*_/*Y*_*novax*_).

The impact of weakening control measures (see *Alternative scenarios – control measures*) is quantified by comparing infection totals between simulations with stronger and weaker control measures. With these quantities,denoted *Y*_*strong*_ and *Y*_*weak*_, respectively, the number of excess infections is given by *Y*_*weak*_ – *Y*_*strong*_. Numbers of infections are also compared between simulations without vaccination (*Y*_*novax*_) and simulations with control measures weakened in multiple ways (*Y*_*multi*_). The difference between these is calculated similarly, as *Y*_*multi*_ – *Y*_*novax*_, but because one is not always larger than the other, we use a diverging color palette to distinguish positive and negative values.

Strain dynamics are also shown in several figures; the number of infections with strain *i*, or *I*_*i*_(*t*), is the sum of all the infected compartments for strain *i* (*I*_*iS*_(*t*), *I*_*iS*(*V*)_(*t*), *I*_*iR*_ (*t*), *I*_*iR*(*V*)_(*t*), *I*_*iV*_(*t*), and *I*_*iV*(*j*)_(*t*)). In one instance, infections are subdivided by the immune status of the host. Infections of naïve individuals with strain *i* are obtained as the sum of compartments *I*_*iS*_(*t*) and *I*_*iS*(*V*)_(*t*); the latter is included because vaccination during infection does not take effect until the infection is cleared. Infections of recovered and immune individuals with strain *i* are obtained by summing the remaining infected compartments.

### Alternative scenarios

Finally, we run through several alternative scenarios which serve two distinct purposes. The purpose of the first group (*Control measures)* is to examine the discrepancy between the default model configuration, which is in many ways a best-case scenario, and more realistic scenarios with various shortcomings in epidemic control measures. The role of the second group (*Model assumptions*) is to determine whether the main findings hold up when key model assumptions are changed.

#### Control measures

The default model configuration, with indefinite continuation of NPIs, complete vaccination coverage, and high vaccine efficacy, represents the best-case scenario in each of these areas; we therefore vary these assumptions to simulate realistic – not pessimistic – shortcomings. We consider three ways in which control measures might suffer (Table 6): lifting NPIs when vaccine coverage reaches 50%, decreasing the final vaccination coverage to 50%, and lowering the peak vaccine efficacy (against WT) to 70%. We repeat simulation sets 5-12 (Table 5) under each of these alternative parameterizations, as well as the combination of all three.

**Table 6.** Alternative scenarios with weakened control measures

| Description | NPIs relaxed | Vaccine coverage ($c$) | Vaccine efficacy ($\omega$) |
| --- | --- | --- | --- |
| Lifting NPIs | At 50% vaccine coverage | 100% | 95% |
| Reduced coverage | No | 50% | 95% |
| Reduced efficacy | No | 100% | 70% |
| Combination | At 50% vaccine coverage | 50% | 70% |

#### Model assumptions

We used three alternative versions of the model to re-run all the simulations listed in Tables 5-6. Each of the alternative models varies one key assumption made in the default model; one can be explained briefly and the other two require more extensive discussion. We therefore consider each alternative model in a different section below.

##### Lower and higher degrees of immune escape

The default model assumes that immune escape manifests as a 40% reduction in cross-reactivity with WT. This value is on the high end of escape estimates for existing variants, but lower degrees of escape almost certainly exist, and higher degrees of immune escape are theoretically possible. We therefore ran simulations with a lower level of immune escape (20%, or 80% cross-reactivity with WT) and a higher level (80% escape, or 20% cross-reactivity). For variants with enhanced transmissibility, we assumed a 60% increase in R_0_, as in the default model. Simulations with a higher level of immune escape were run for an extended duration (six years) because the occurrence of a second wave of variant infections in some scenarios increased the time frame over which epidemic dynamics were occurring.

##### Rolling lockdowns

In the default model, we assume that nonpharmaceutical interventions (NPIs) are fixed at an intensity that reduces transmission by 40% (Table 3). This configuration generates dynamics that are easy to understand and analyze, but is not broadly representative of the dynamics of the COVID-19 pandemic. In many parts of the world, relatively weak NPIs are periodically supplanted by more stringent measures when case numbers grow too large; after a period of sustained decline, the more severe control measures are turned off again. We use an alternative model configuration to simulate this strategy, which we refer to as “rolling lockdowns.”

In the alternative configuration, NPIs switch between two different intensities, which reduce transmission by 30% and 70%, respectively. The high-intensity control measures are triggered when the number of current infections – lagged by 14 days to simulate various delays between infection and reporting – exceeds 1% of the total population. Control measures revert to the lower intensity when the number of infections (lagged by 14 days) drops below the threshold again. Shifts between low and high intensity of NPIs cannot occur less than 14 days apart (the length of the reporting lag), to ensure that shift *n* is precipitated only by events following shift *n* – 1.

##### All-or-nothing immunity

The default model configuration assumes that partial cross-protection from infection or partial immunity from vaccination is imperfect at the individual level, meaning the probability of infection given exposure is reduced but not eliminated, and the degree of protection is the same for all individuals. This is sometimes called “leaky” immunity; an alternative formulation, termed “all-or-nothing” immunity, does not accommodate partial protection for individuals. Upon infection or vaccination, some individuals acquire complete, sterilizing protection, and others acquire no protection at all.

We use an alternative version of the ODE model described above to re-run the same scenarios with all-or-nothing immunity instead of leaky immunity. Although the names of the state variables are largely unchanged, many are defined differently (Table 7). In the default model, states are defined by history of infection and vaccination (Table 2); in the alternative model, states are defined in terms of strain-specific immunity, regardless of how this immunity was acquired. For example, individuals in compartment *V*_*i*_ are vaccinated and immune to strain *i*, but the compartment is not specific to a single history, as it encompasses individuals with and without prior infection by strain *i* (Fig 7).

**Table 7.** State variables for alternative model with all-or-nothing immunity

| State variable | Definition |
| --- | --- |
| $S$ | Susceptible |
| $I_{iS}$ | Infected with strain $i$, previously susceptible |
| $I_{iS(V)}$ | Infected with strain $i$, previously susceptible, vaccinated during current infection |
| $R_i$ | Recovered and immune to strain $i$ |
| $R_{ij}$ | Recovered and immune to both strain $i$ and strain $j$ |
| $I_{iR}$ | Infected with strain $i$, previously recovered (from strain $j$) |
| $I_{iR(V)}$ | Infected with strain $i$, previously recovered (from strain $j$), vaccinated during current infection |
| $V_i$ | Vaccinated and immune to strain $i$ |
| $V_{ij}$ | Vaccinated and immune to both strain $i$ and strain $j$ |
| $V_0$ | Vaccinated but not immune to either strain |
| $I_{iV(j)}$ | Previously vaccinated, immune to strain $j$, currently infected with strain $i$ |
| $I_{iV(0)}$ | Previously vaccinated, not immune to either strain, currently infected with strain $i$ |

**Fig 7.**
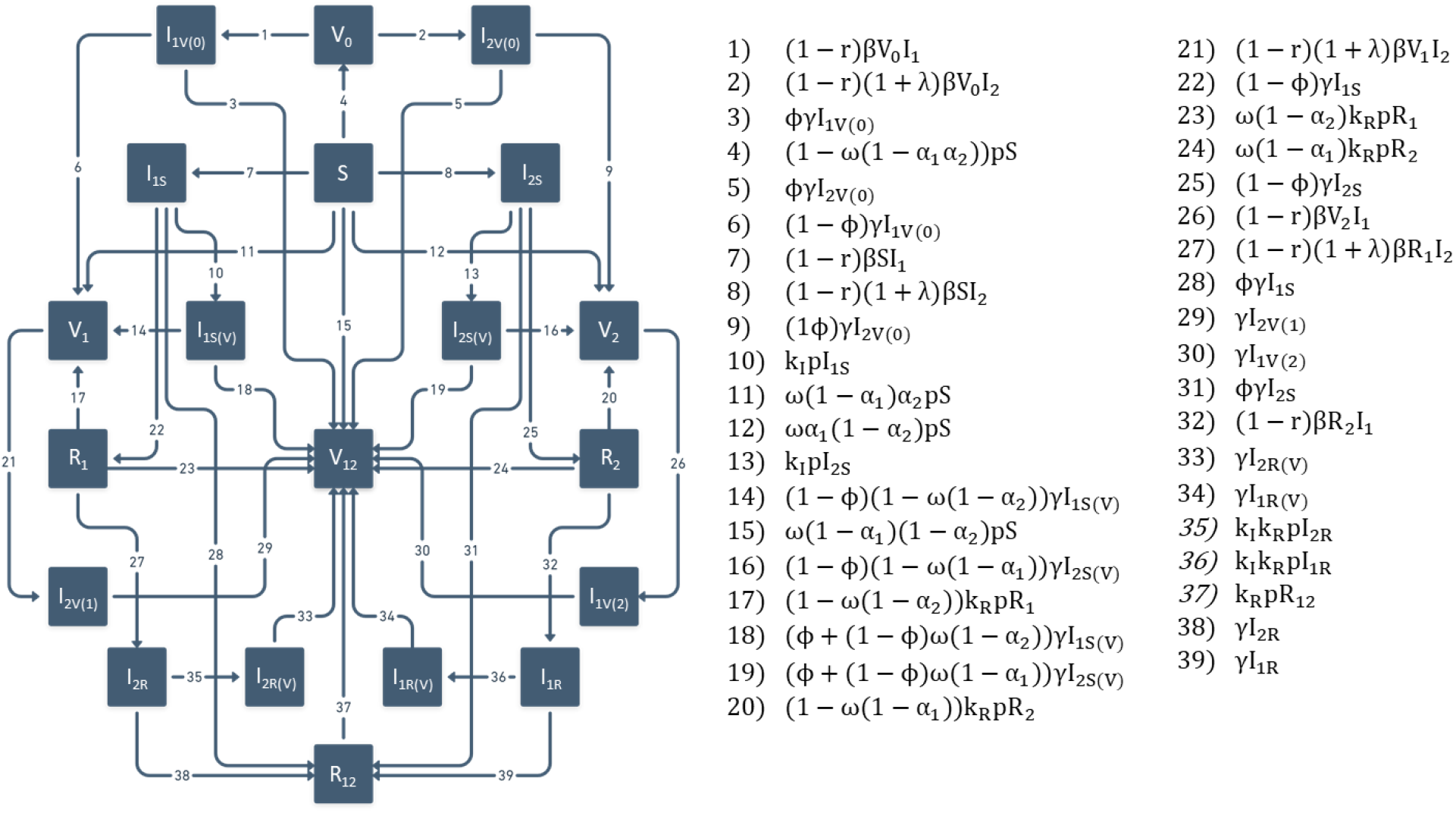
Diagram and rates of movement between compartments for alternative model with all-or-nothing immunity.

The model parameters are generally the same, except that parameters relating to partial immunity (*ϕ, ω, α*_1_, and *α*_2_) are defined in terms of probabilities of protection, rather than degrees of protection (Table 8). Rates of movement between compartments are very different from the default model (Fig 7), as are the differential equations (Eqs 25-44).

**Table 8.** Parameters for alternative model with all-or-nothing immunity

| Parameter | Definition | Default value |
| --- | --- | --- |
| $N$ | Population size | $10^8$ |
| $R_0$ | Reproduction number (WT) | 2.5 |
| $\gamma$ | Recovery rate | $1/14 \text{ days}^{-1}$ |
| $\beta$ | Transmission rate | $\frac{R_0 \gamma}{N}$ |
| $\phi$ | Probability of cross-protection given infection with either strain | See Tables 4-5 |
| $\omega$ | Maximum vaccine efficacy (probability of protection against perfect antigenic match) | 0.95 |
| $\alpha_1$ | Probability vaccine fails to elicit immunity against strain 1 (due to antigenic mismatch) | 0 |
| $\alpha_2$ | Probability vaccine fails to elicit immunity against strain 2 (due to antigenic mismatch) | $1 - \phi$ |
| $\kappa_I$ | Eligibility of infected individuals for vaccination; 1 = eligible, 0 = ineligible | 1 |
| $\kappa_R$ | Eligibility of recovered individuals for vaccination; 1 = eligible, 0 = ineligible | 1 |
| $\lambda$ | Increase in transmissibility of variant relative to WT | See Tables 4-5 |
| $\tau_2$ | Time of variant introduction | 9 months |
| $\tau_{vax}$ | Start of vaccine rollout | Varies |
| $v$ | Daily per-capita vaccination rate | Varies |
| $c$ | Final vaccination coverage | 100% |
| $r$ | Reduction in transmission due to nonpharmaceutical interventions | 0.4 |

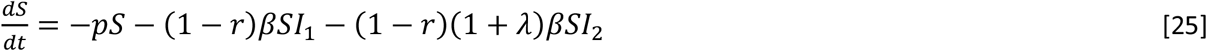

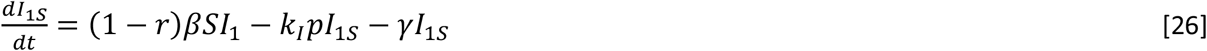

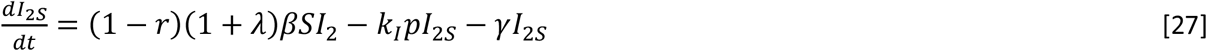

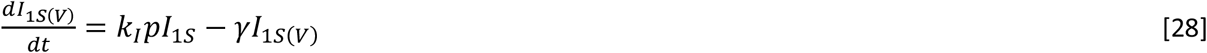

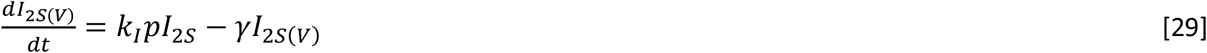

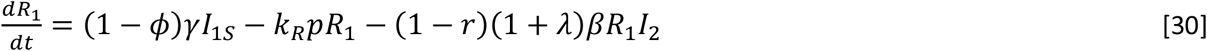

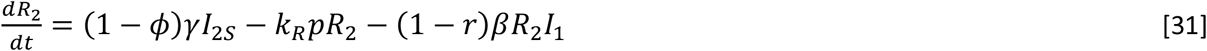

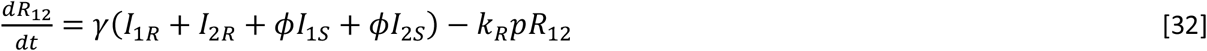

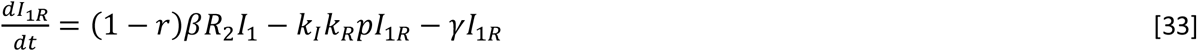

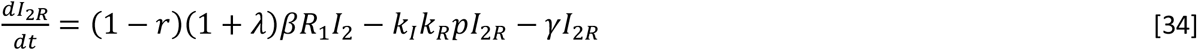

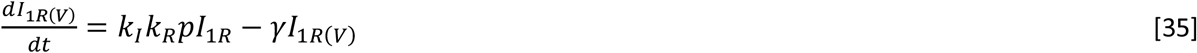

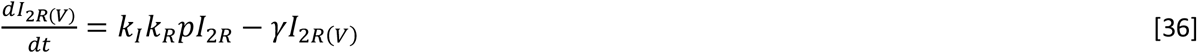

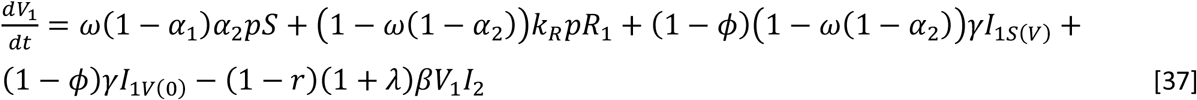

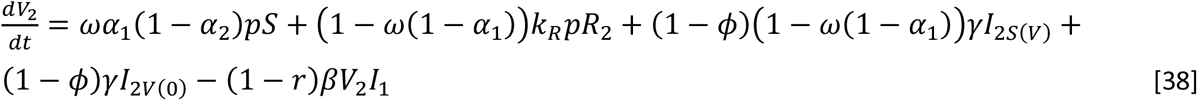

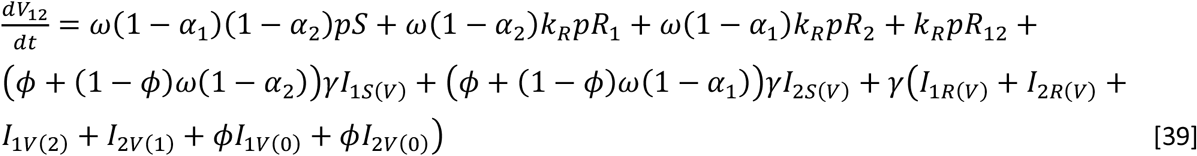

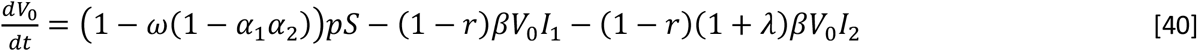

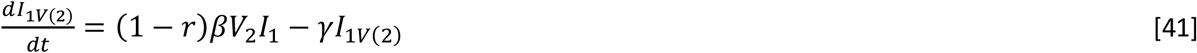

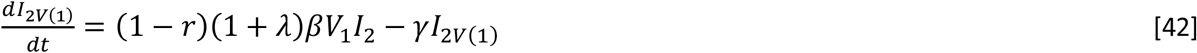

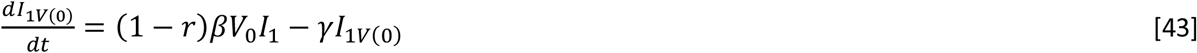

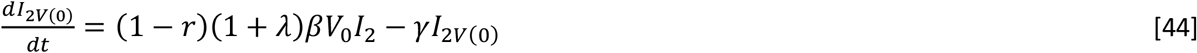

### Software

The model was implemented in R (version 4.1.0). The code and saved model output can be downloaded from Open Science Framework at https://osf.io/z9x2p/?view_only=6def9d71316645d0a54a30fa6e9ffe13.

## Data Availability

Model code is freely available at https://osf.io/z9x2p/?view_only=6def9d71316645d0a54a30fa6e9ffe13

## References

1. Weekly epidemiological update on COVID-19 - 10 August 2021. 52 ed: World Health Organization.

2. Davies NG, Abbott S, Barnard RC, Jarvis CI, Kucharski AJ, Munday JD, et al. Estimated transmissibility and impact of SARS-CoV-2 lineage B.1.1.7 in England. Science. 2021;372(6538). doi: 10.1126/science.abg3055.

3. Volz E, Mishra S, Chand M, Barrett JC, Johnson R, Geidelberg L, et al. Assessing transmissibility of SARS-CoV-2 lineage B.1.1.7 in England. Nature. 2021;593(7858):266–9. doi: 10.1038/s41586-021-03470-x.

4. Planas D, Bruel T, Grzelak L, Guivel-Benhassine F, Staropoli I, Porrot F, et al. Sensitivity of infectious SARS-CoV-2 B.1.1.7 and B.1.351 variants to neutralizing antibodies. Nat Med. 2021;27(5):917–24. doi: 10.1038/s41591-021-01318-5.

5. Supasa P, Zhou D, Dejnirattisai W, Liu C, Mentzer AJ, Ginn HM, et al. Reduced neutralization of SARS-CoV-2 B.1.1.7 variant by convalescent and vaccine sera. Cell. 2021. doi: 10.1016/j.cell.2021.02.033.

6. Wang P, Nair MS, Liu L, Iketani S, Luo Y, Guo Y, et al. Antibody Resistance of SARS-CoV-2 Variants B.1.351 and B.1.1.7. Nature. 2021. doi: 10.1038/s41586-021-03398-2.

7. Graham MS, Sudre CH, May A, Antonelli M, Murray B, Varsavsky T, et al. Changes in symptomatology, reinfection, and transmissibility associated with the SARS-CoV-2 variant B.1.1.7: an ecological study. Lancet Public Health. 2021;6(5):e335–e45. doi: 10.1016/s2468-2667(21)00055-4.

8. Tegally H, Wilkinson E, Giovanetti M, Iranzadeh A, Fonseca V, Giandhari J, et al. Detection of a SARS-CoV-2 variant of concern in South Africa. Nature. 2021;592(7854):438–43. doi: 10.1038/s41586-021-03402-9.

9. Cele S, Gazy I, Jackson L, Hwa SH, Tegally H, Lustig G, et al. Escape of SARS-CoV-2 501Y.V2 from neutralization by convalescent plasma. Nature. 2021;593(7857):142–6. doi: 10.1038/s41586-021-03471-w.

10. Wibmer CK, Ayres F, Hermanus T, Madzivhandila M, Kgagudi P, Oosthuysen B, et al. SARS-CoV-2 501Y.V2 escapes neutralization by South African COVID-19 donor plasma. bioRxiv. 2021. doi: 10.1101/2021.01.18.427166.

11. Zhou D, Dejnirattisai W, Supasa P, Liu C, Mentzer AJ, Ginn HM, et al. Evidence of escape of SARS-CoV-2 variant B.1.351 from natural and vaccine-induced sera. Cell. 2021. doi: 10.1016/j.cell.2021.02.037.

12. Geers D, Shamier MC, Bogers S, den Hartog G, Gommers L, Nieuwkoop NN, et al. SARS-CoV-2 variants of concern partially escape humoral but not T-cell responses in COVID-19 convalescent donors and vaccinees. Science Immunology. 2021;6(59):eabj1750. doi: 10.1126/sciimmunol.abj1750.

13. Tarke A, Sidney J, Methot N, Yu ED, Zhang Y, Dan JM, et al. Impact of SARS-CoV-2 variants on the total CD4(+) and CD8(+) T cell reactivity in infected or vaccinated individuals. Cell Rep Med. 2021:100355. doi: 10.1016/j.xcrm.2021.100355.

14. Faria NR, Mellan TA, Whittaker C, Claro IM, Candido DdS, Mishra S, et al. Genomics and epidemiology of the P.1 SARS-CoV-2 lineage in Manaus, Brazil. Science. 2021;372(6544):815–21. doi: 10.1126/science.abh2644.

15. Wang P, Casner RG, Nair MS, Wang M, Yu J, Cerutti G, et al. Increased resistance of SARS-CoV-2 variant P.1 to antibody neutralization. Cell Host Microbe. 2021;29(5):747-51.e4. doi: 10.1016/j.chom.2021.04.007.

16. Dagpunar J. Interim estimates of increased transmissibility, growth rate, and reproduction number of the Covid-19 B.1.617.2 variant of concern in the United Kingdom. medRxiv. 2021:2021.06.03.21258293. doi: 10.1101/2021.06.03.21258293.

17. Allen H, Vusirikala A, Flannagan J, Twohig K, Groves N, Lopez-Bernal J, et al. Increased household transmission of COVID-19 cases associated with SARS-CoV-2 Variant of Concern B.1.617.2: a national case-control study. 2021.

18. Hoffmann M, Hofmann-Winkler H, Krüger N, Kempf A, Nehlmeier I, Graichen L, et al. SARS-CoV-2 variant B.1.617 is resistant to bamlanivimab and evades antibodies induced by infection and vaccination. Cell Reports. 2021;36(3):109415. doi: https://doi.org/10.1016/j.celrep.2021.109415.

19. Planas D, Veyer D, Baidaliuk A, Staropoli I, Guivel-Benhassine F, Rajah MM, et al. Reduced sensitivity of SARS-CoV-2 variant Delta to antibody neutralization. Nature. 2021;596(7871):276–80. doi: 10.1038/s41586-021-03777-9.

20. Lopez Bernal J, Andrews N, Gower C, Gallagher E, Simmons R, Thelwall S, et al. Effectiveness of Covid-19 Vaccines against the B.1.617.2 (Delta) Variant. N Engl J Med. 2021;385(7):585–94. doi: 10.1056/NEJMoa2108891.

21. Mathieu E, Ritchie H, Ortiz-Ospina E, Roser M, Hasell J, Appel C, et al. A global database of COVID-19 vaccinations. Nature Human Behaviour. 2021;5(7):947–53. doi: 10.1038/s41562-021-01122-8.

22. Sadoff J, Gray G, Vandebosch A, Cardenas V, Shukarev G, Grinsztejn B, et al. Safety and Efficacy of Single-Dose Ad26.COV2.S Vaccine against Covid-19. New England Journal of Medicine.15. doi: 10.1056/NEJMoa2101544.

23. Haas EJ, Angulo FJ, McLaughlin JM, Anis E, Singer SR, Khan F, et al. Impact and effectiveness of mRNA BNT162b2 vaccine against SARS-CoV-2 infections and COVID-19 cases, hospitalisations, and deaths following a nationwide vaccination campaign in Israel: an observational study using national surveillance data. Lancet. 2021;397(10287):1819–29. doi: 10.1016/s0140-6736(21)00947-8.

24. Abu-Raddad LJ, Chemaitelly H, Butt AA. Effectiveness of the BNT162b2 Covid-19 Vaccine against the B.1.1.7 and B.1.351 Variants. New England Journal of Medicine. 2021. doi: 10.1056/NEJMc2104974.

25. Baden LR, El Sahly HM, Essink B, Kotloff K, Frey S, Novak R, et al. Efficacy and Safety of the mRNA-1273 SARS-CoV-2 Vaccine. New England Journal of Medicine. 2021;384(5):403–16. doi: 10.1056/NEJMoa2035389.

26. Polack FP, Thomas SJ, Kitchin N, Absalon J, Gurtman A, Lockhart S, et al. Safety and Efficacy of the BNT162b2 mRNA Covid-19 Vaccine. New England Journal of Medicine. 2020;383(27):2603–15. doi: 10.1056/NEJMoa2034577.

27. Voysey M, Clemens SAC, Madhi SA, Weckx LY, Folegatti PM, Aley PK, et al. Safety and efficacy of the ChAdOx1 nCoV-19 vaccine (AZD1222) against SARS-CoV-2: an interim analysis of four randomised controlled trials in Brazil, South Africa, and the UK. Lancet. 2021;397(10269):99–111. doi: 10.1016/s0140-6736(20)32661-1.

28. Collier DA, De Marco A, Ferreira I, Meng B, Datir RP, Walls AC, et al. Sensitivity of SARS-CoV-2 B.1.1.7 to mRNA vaccine-elicited antibodies. Nature. 2021;593(7857):136–41. doi: 10.1038/s41586-021-03412-7.

29. Muik A, Wallisch AK, Sänger B, Swanson KA, Mühl J, Chen W, et al. Neutralization of SARS-CoV-2 lineage B.1.1.7 pseudovirus by BNT162b2 vaccine-elicited human sera. Science. 2021;371(6534):1152–3. doi: 10.1126/science.abg6105.

30. Emary KRW, Golubchik T, Aley PK, Ariani CV, Angus B, Bibi S, et al. Efficacy of ChAdOx1 nCoV-19 (AZD1222) vaccine against SARS-CoV-2 variant of concern 202012/01 (B.1.1.7): an exploratory analysis of a randomised controlled trial. Lancet. 2021;397(10282):1351–62. doi: 10.1016/s0140-6736(21)00628-0.

31. Kustin T, Harel N, Finkel U, Perchik S, Harari S, Tahor M, et al. Evidence for increased breakthrough rates of SARS-CoV-2 variants of concern in BNT162b2 mRNA vaccinated individuals. medRxiv.2021:2021.04.06.21254882. doi: 10.1101/2021.04.06.21254882.

32. Becker M, Dulovic A, Junker D, Ruetalo N, Kaiser PD, Pinilla YT, et al. Immune response to SARS-CoV-2 variants of concern in vaccinated individuals. Nat Commun. 2021;12(1):3109. doi: 10.1038/s41467-021-23473-6.

33. Garcia-Beltran WF, Lam EC, St Denis K, Nitido AD, Garcia ZH, Hauser BM, et al. Multiple SARS-CoV-2 variants escape neutralization by vaccine-induced humoral immunity. Cell. 2021. doi: 10.1016/j.cell.2021.03.013.

34. Lustig Y, Zuckerman N, Nemet I, Atari N, Kliker L, Regev-Yochay G, et al. Neutralising capacity against Delta (B.1.617.2) and other variants of concern following Comirnaty (BNT162b2, BioNTech/Pfizer) vaccination in health care workers, Israel. Euro Surveill. 2021;26(26). doi: 10.2807/1560-7917.Es.2021.26.26.2100557.

35. Madhi SA, Baillie V, Cutland CL, Voysey M, Koen AL, Fairlie L, et al. Efficacy of the ChAdOx1 nCoV-19 Covid-19 Vaccine against the B.1.351 Variant. N Engl J Med. 2021. doi: 10.1056/NEJMoa2102214.

36. Mor O, Zuckerman NS, Hazan I, Fluss R, Ash N, Ginish N, et al. BNT162b2 Vaccination efficacy is marginally affected by the SARS-CoV-2 B.1.351 variant in fully vaccinated individuals. medRxiv. 2021:2021.07.20.21260833. doi: 10.1101/2021.07.20.21260833.

37. Dejnirattisai W, Zhou D, Supasa P, Liu C, Mentzer AJ, Ginn HM, et al. Antibody evasion by the P.1 strain of SARS-CoV-2. Cell. 2021;184(11):2939-54.e9. doi: https://doi.org/10.1016/j.cell.2021.03.055.

38. Liu C, Ginn HM, Dejnirattisai W, Supasa P, Wang B, Tuekprakhon A, et al. Reduced neutralization of SARS-CoV-2 B.1.617 by vaccine and convalescent serum. Cell. 2021;184(16):4220-36.e13. doi: 10.1016/j.cell.2021.06.020.

39. Kennedy DA, Read AF. Why does drug resistance readily evolve but vaccine resistance does not? Proc Biol Sci. 2017;284(1851). doi: 10.1098/rspb.2016.2562.

40. Ashby B, Thompson RN. Non-pharmaceutical interventions and the emergence of pathogen variants. medRxiv. 2021:2021.05.27.21257938. doi: 10.1101/2021.05.27.21257938.

41. Sheikh A, McMenamin J, Taylor B, Robertson C. SARS-CoV-2 Delta VOC in Scotland: demographics, risk of hospital admission, and vaccine effectiveness. Lancet. 2021;397(10293):2461–2. doi: 10.1016/s0140-6736(21)01358-1.

42. Ong SWX, Chiew CJ, Ang LW, Mak TM, Cui L, Toh M, et al. Clinical and virological features of SARS-CoV-2 variants of concern: a retrospective cohort study comparing B.1.1.7 (Alpha), B.1.315 (Beta), and B.1.617.2 (Delta). Clin Infect Dis. 2021. doi: 10.1093/cid/ciab721.

