## Supplementary material for "Population impact of SARS-CoV-2 variants with enhanced transmissibility and/or partial immune escape": Figs S1-S18

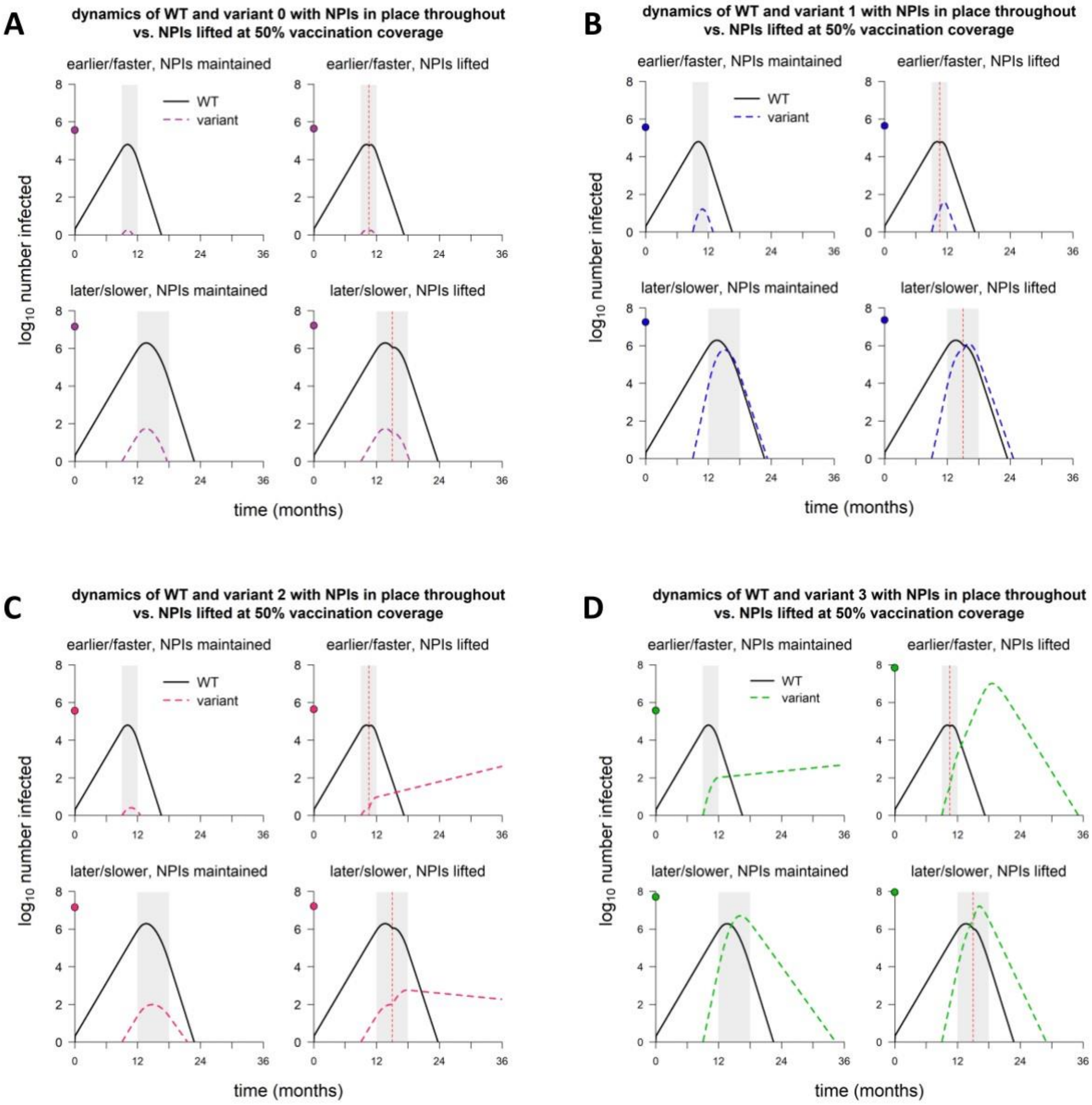


**Fig S1.** Dynamics of WT and variant strains with rapid vs. delayed vaccine rollout and continuation vs. discontinuation of nonpharmaceutical interventions. (A) Dynamics of WT and variant 0. (B) WT and variant 1. (C) WT and variant 2. (D) WT and variant 3. Simulation conditions as follows: NPIs maintained = in place throughout; NPIs lifted = discontinued when vaccine coverage reaches 50%; earlier/faster vaccine rollout = starting at 9 months and lasting 3 months; later/slower rollout = starting at 12 months and lasting 6 months. Variant introduced at 9 months in all simulations. Solid/black lines, WT; colored/dashed lines, variants; gray shading, vaccine rollout; vertical dashed line, 50% vaccination coverage reached and NPIs lifted. In each panel, point on y-axis indicates total number of infections over the entire simulation. Variant phenotypes are as follows: variant 0, identical to WT; variant 1, 60% greater transmissibility; variant 2, 40% immune escape; variant 3, 60% greater transmissibility and 40% immune escape.

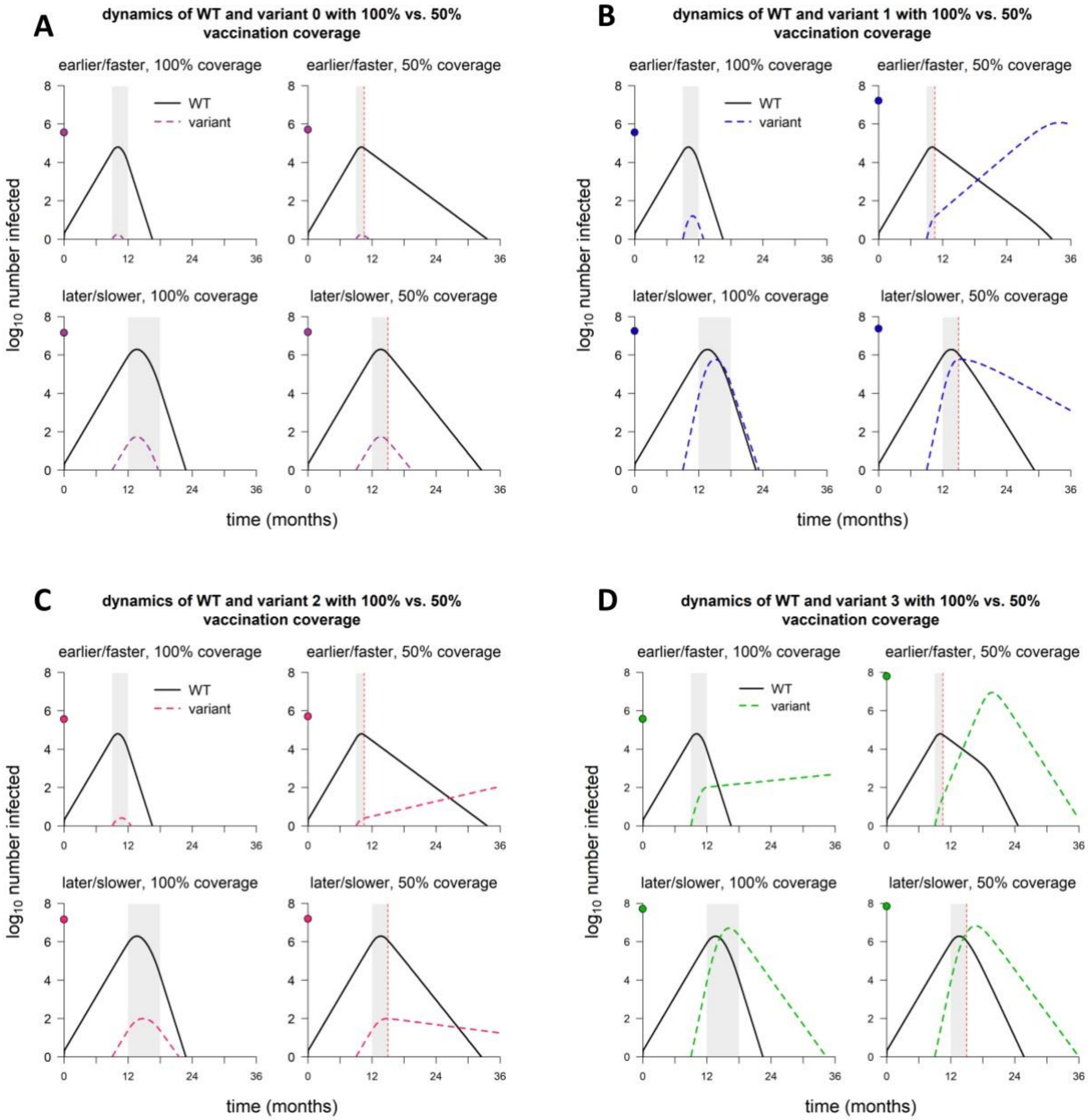


**Fig S2.** Dynamics of WT and variant strains with rapid vs. delayed vaccine rollout and 100% vaccination coverage vs. 50% coverage. (A) Dynamics of WT and variant 0. (B) WT and variant 1. (C) WT and variant 2. (D) WT and variant 3. Simulation conditions as follows: earlier/faster vaccine rollout = starting at 9 months and lasting 3 months (100% coverage) or 1.5 months (50% coverage); later/slower rollout = starting at 12 months and lasting 6 months (100% coverage) or 3 months (50% coverage). Variant introduced at 9 months in all simulations. Solid/black lines, WT; colored/dashed lines, variants; gray shading, vaccine rollout; vertical dashed line, 50% vaccination coverage reached. In each panel, point on y-axis indicates total number of infections over the entire simulation. Variant phenotypes are as follows: variant 0, identical to WT; variant 1, 60% greater transmissibility; variant 2, 40% immune escape; variant 3, 60% greater transmissibility and 40% immune escape.

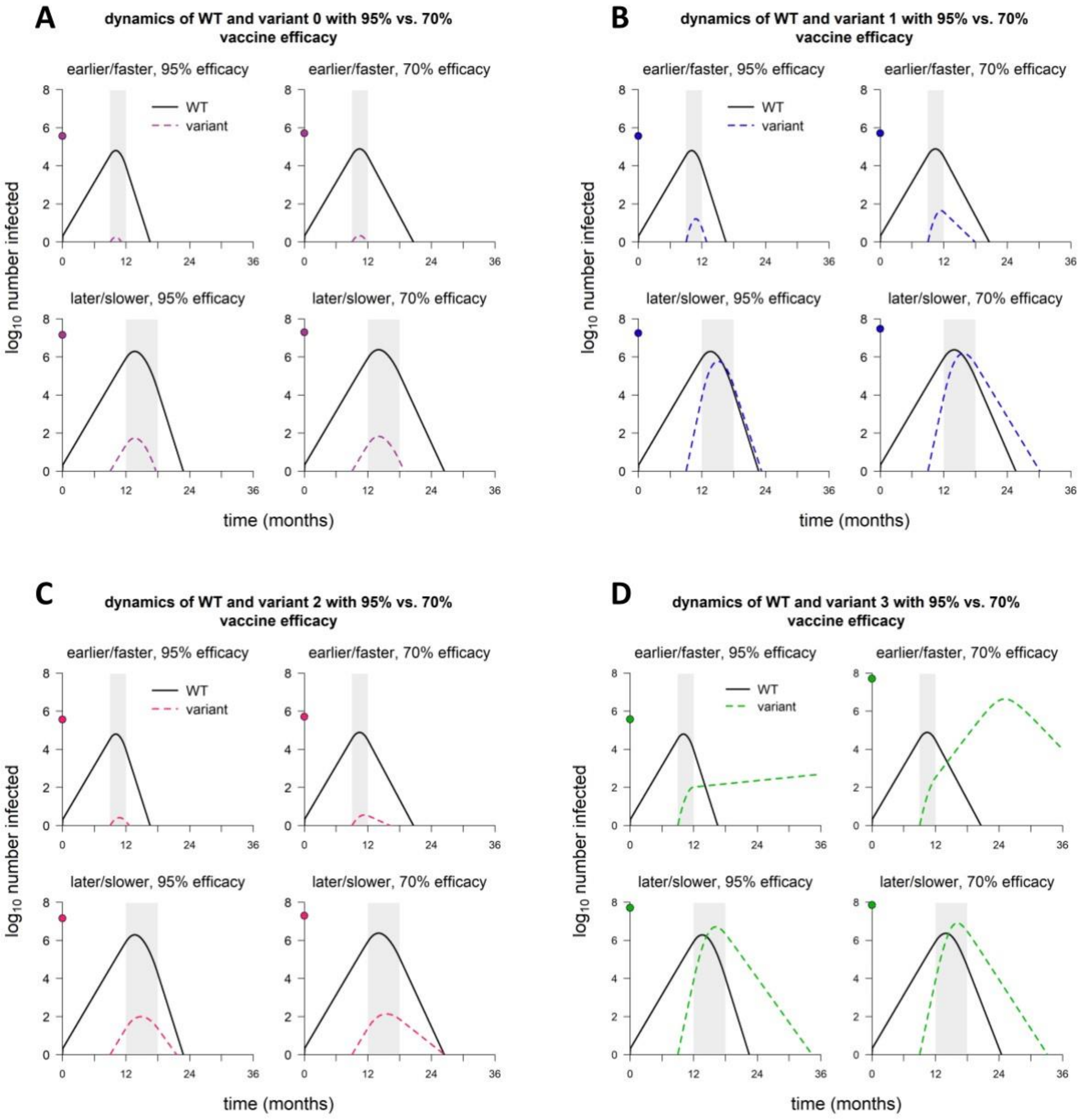


**Fig S3.** Dynamics of WT and variant strains with rapid vs. delayed vaccine rollout and 95% baseline vaccine efficacy vs. 70% baseline efficacy. (A) Dynamics of WT and variant 0. (B) WT and variant 1. (C) WT and variant 2. (D) WT and variant 3. Simulation conditions as follows: earlier/faster vaccine rollout = starting at 9 months and lasting 3 months; later/slower rollout = starting at 12 months and lasting 6 months. Variant introduced at 9 months in all simulations. Solid/black lines, WT; colored/dashed lines, variants; gray shading, vaccine rollout. In each panel, point on y-axis indicates total number of infections over the entire simulation. Variant phenotypes are as follows: variant 0, identical to WT; variant 1, 60% greater transmissibility; variant 2, 40% immune escape; variant 3, 60% greater transmissibility and 40% immune escape.

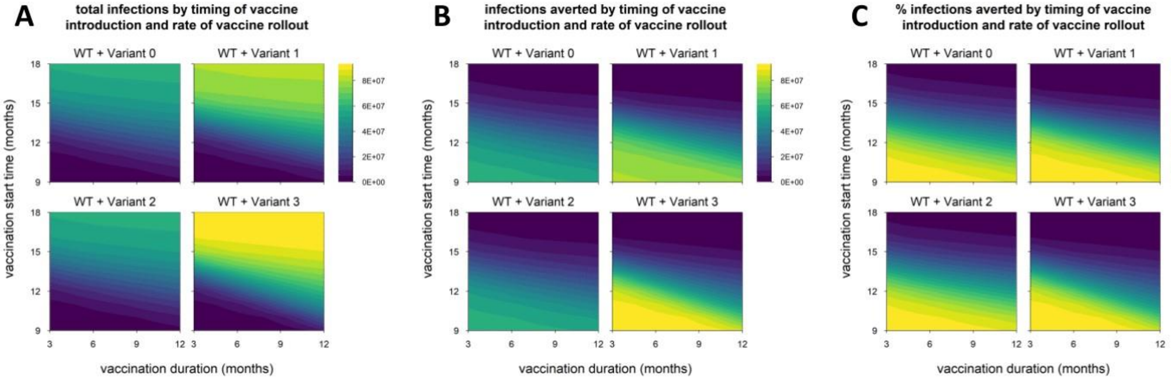


**Fig S4.** Epidemic size and vaccination impact for alternative model with lower degree of immune escape. (A) Total infections (WT + variant) in simulations with each hypothetical variant, for varying rates of vaccination (vaccination duration, x-axis) and time of vaccine introduction (vaccination start time, y-axis); shaded contours represent total infections. (B) Number of infections (WT + variant) averted by vaccination; shading represents number of infections averted. (C) Percentage of infections (WT + variant) averted by vaccination; shading represents % infections averted. Variant introduced at 9 months in all simulations. Variant phenotypes are as follows: variant 0, identical to WT; variant 1, 60% greater transmissibility; variant 2, 20% immune escape; variant 3, 60% greater transmissibility and 20% immune escape.

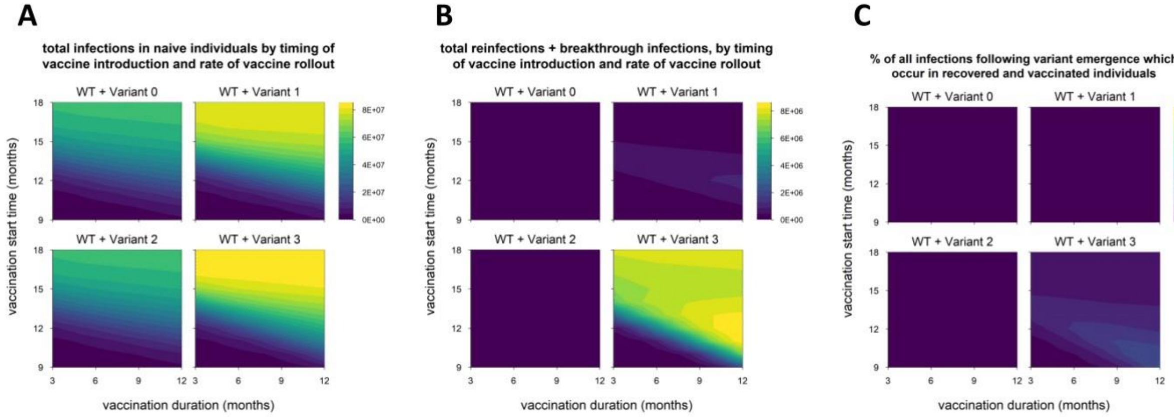


**Fig S5.** Breakdown of infections by immune status (naïve vs. recovered/vaccinated) for alternative model with lower degree of immune escape. (A) Total infections in naïve individuals, in simulations with each variant, for varying rates of vaccination (x-axis) and time of vaccine introduction (y-axis); shaded contours represent total infections. (B) Total infections in recovered and vaccinated individuals (reinfections and breakthrough infections); shaded contours represent total infections. (C) Percentage of all infections occurring in recovered/vaccinated individuals (reinfections/breakthrough infections), starting from the time of variant emergence; shading represents percentage of infections occurring in recovered/vaccinated individuals. Variant introduced at 9 months in all simulations. Variant phenotypes are as follows: variant 0, identical to WT; variant 1, 60% greater transmissibility; variant 2, 20% immune escape; variant 3, 60% greater transmissibility and 20% immune escape.

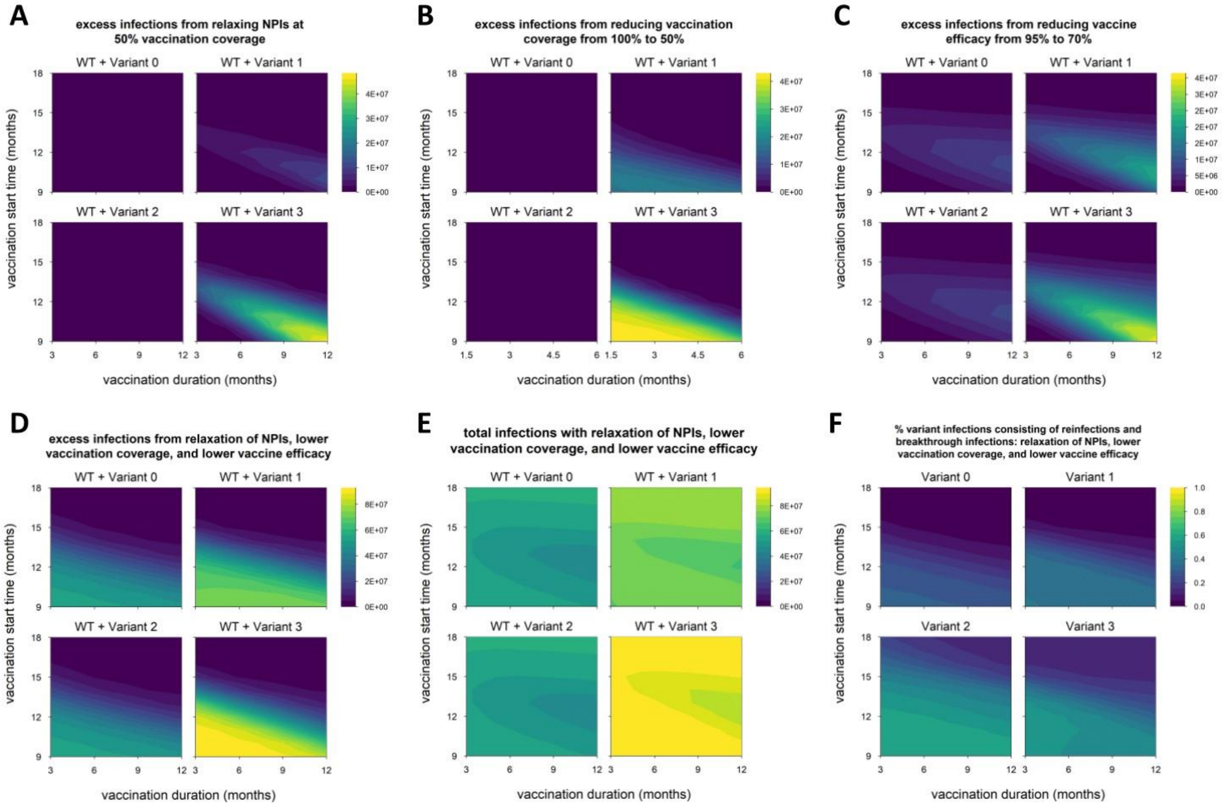


**Fig S6**. Impact of reduced control measures on epidemic size for alternative model with lower degree of immune escape. (A) Excess infections from lifting nonpharmaceutical interventions (NPIs) when vaccination coverage reaches 50% (default condition is NPIs continued indefinitely); shading represents excess infections compared to default conditions/parameters. (B) Excess infections from reducing vaccination coverage from 100% to 50%. (C) Excess infections from reducing baseline vaccine efficacy (against WT) from 95% to 70%. (D) Excess infections from the combination of conditions A through C (lifting NPIs, reduced coverage and reduced efficacy). (E) Total infections (WT + variant) under the combined conditions of panels A through C; shading represent total infections. (F) Percentage of variant infections composed of reinfections and breakthrough infections under the combined conditions of panels A through C; shading represent percentage of variant infections occurring in recovered/vaccinated individuals. Variant introduced at 9 months in all simulations. Variant phenotypes are as follows: variant 0, identical to WT; variant 1, 60% greater transmissibility; variant 2, 20% immune escape; variant 3, 60% greater transmissibility and 20% immune escape.

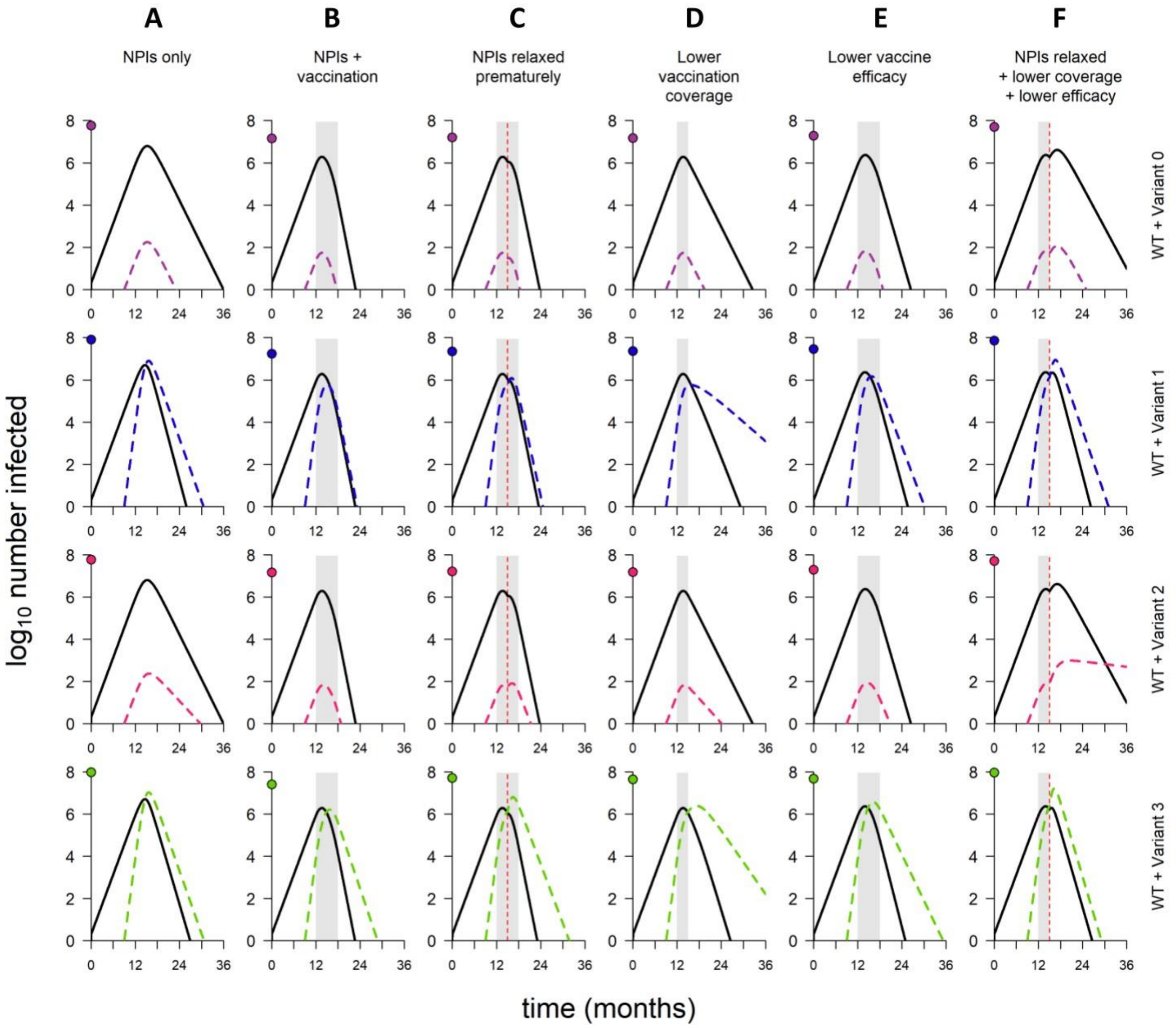


**Fig S7.** Dynamics of WT and variants, in simulations with varying combinations of control measures, for alternative model with lower degree of immune escape. (A) No vaccination but NPIs in place throughout. (B) Default model conditions (NPIs in place throughout, 100% vaccination coverage, 95% vaccine efficacy against WT). (C) NPIs lifted when vaccination coverage reaches 50% (other model conditions as in B). (D) 50% vaccination coverage (other model conditions as in B). (E) 70% vaccine efficacy (other model conditions as in B). (F) Combination of conditions C through E (NPIs relaxed, 50% vaccination coverage, and 70% vaccine efficacy). In each panel, point on y-axis indicates the total number of infections over the entire simulation. Variant introduced at 9 months in all simulations; in simulations with vaccination, rollout begins at 12 months and lasts 6 months if final coverage is 100%, 3 months if final coverage is 50%. Solid/black lines, WT; colored/dashed lines, variants; gray shading, vaccine rollout; dashed vertical line, 50% vaccination coverage. Variant phenotypes are as follows: variant 0, identical to WT; variant 1, 60% greater transmissibility; variant 2, 20% immune escape; variant 3, 60% greater transmissibility and 20% immune escape.

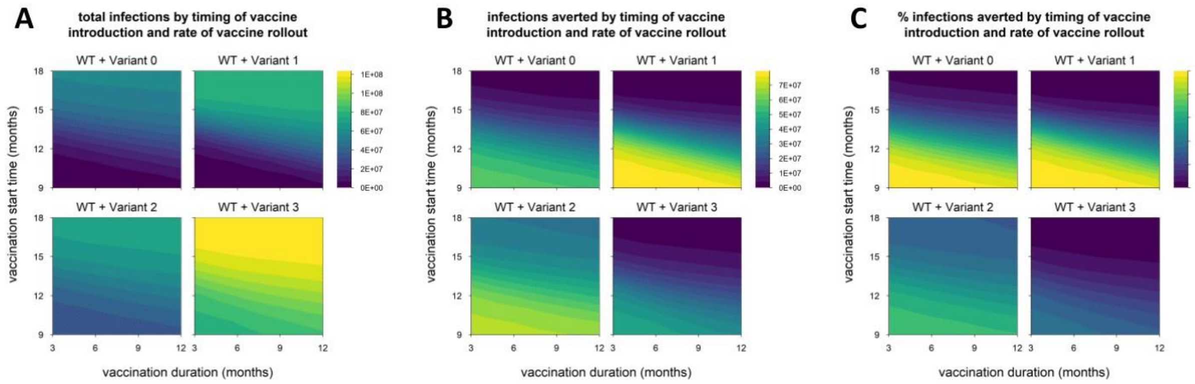


**Fig S8.** Epidemic size and vaccination impact for alternative model with higher degree of immune escape. (A) Total infections (WT + variant) in simulations with each hypothetical variant, for varying rates of vaccination (vaccination duration, x-axis) and time of vaccine introduction (vaccination start time, y-axis); shaded contours represent total infections. (B) Number of infections (WT + variant) averted by vaccination; shading represents number of infections averted. (C) Percentage of infections (WT + variant) averted by vaccination; shading represents % infections averted. All simulations run for an extended duration of six years (default condition is three years); variant introduced at 9 months in all simulations. Variant phenotypes are as follows: variant 0, identical to WT; variant 1, 60% greater transmissibility; variant 2, 80% immune escape; variant 3, 60% greater transmissibility and 80% immune escape.

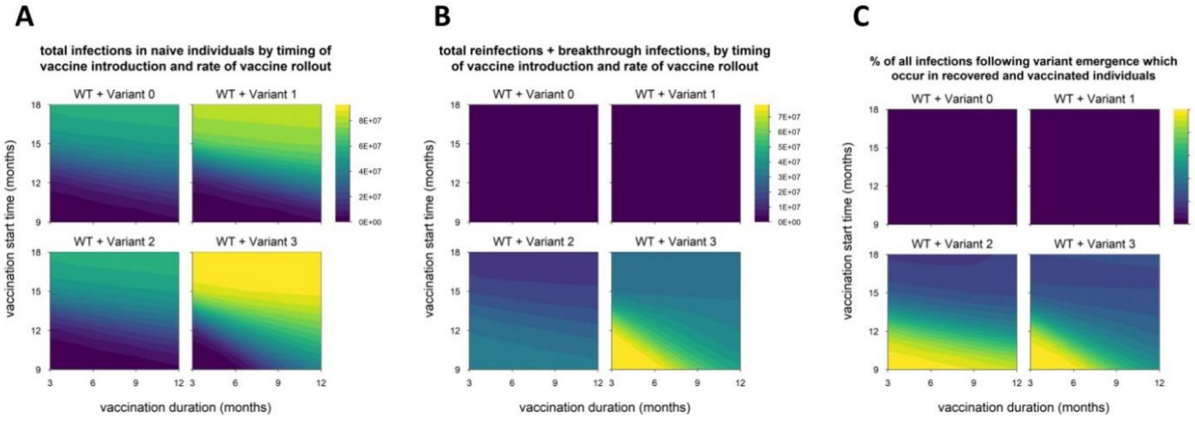


**Fig S9.** Breakdown of infections by immune status (naïve vs. recovered/vaccinated) for alternative model with higher degree of immune escape. (A) Total infections in naïve individuals, in simulations with each variant, for varying rates of vaccination (x-axis) and time of vaccine introduction (y-axis); shaded contours represent total infections. (B) Total infections in recovered and vaccinated individuals (reinfections and breakthrough infections); shaded contours represent total infections. (C) Percentage of all infections occurring in recovered/vaccinated individuals (reinfections/breakthrough infections), starting from the time of variant emergence; shading represents percentage of infections occurring in recovered/vaccinated individuals. All simulations run for an extended duration of six years (default condition is three years); variant introduced at 9 months in all simulations.Variant phenotypes are as follows: variant 0, identical to WT; variant 1, 60% greater transmissibility; variant 2, 80% immune escape; variant 3, 60% greater transmissibility and 80% immune escape.

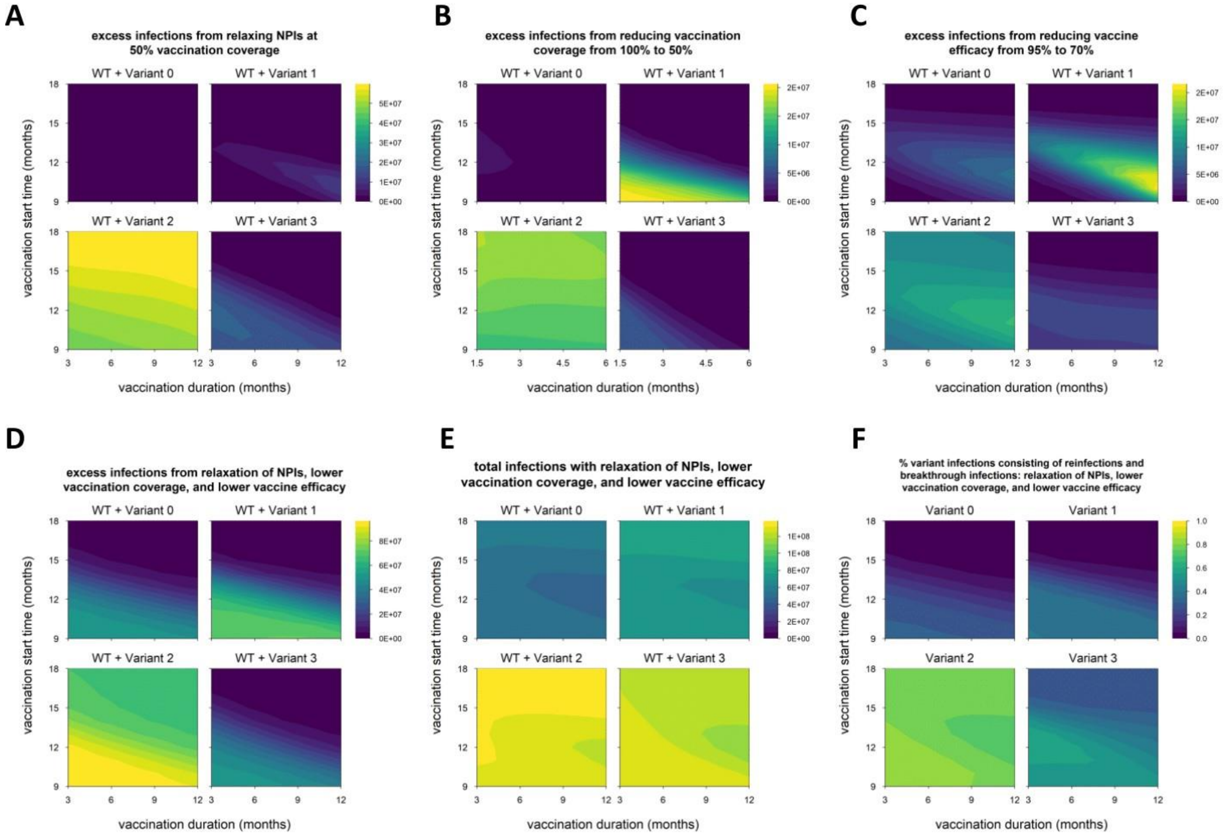


**Fig S10.** Impact of reduced control measures on epidemic size for alternative model with higher degree of immune escape. (A) Excess infections from lifting nonpharmaceutical interventions (NPIs) when vaccination coverage reaches 50% (default condition is NPIs continued indefinitely); shading represents excess infections compared to default conditions/parameters. (B) Excess infections from reducing vaccination coverage from 100% to 50%. (C) Excess infections from reducing baseline vaccine efficacy (against WT) from 95% to 70%. (D) Excess infections from the combination of conditions A through C (lifting NPIs, reduced coverage and reduced efficacy). (E) Total infections (WT + variant) under the combined conditions of panels A through C; shading represent total infections. (F) Percentage of variant infections composed of reinfections and breakthrough infections under the combined conditions of panels A through C; shading represent percentage of variant infections occurring in recovered/vaccinated individuals. All simulations run for an extended duration of six years (default condition is three years); variant introduced at 9 months in all simulations. Variant phenotypes are as follows: variant 0, identical to WT; variant 1, 60% greater transmissibility; variant 2, 80% immune escape; variant 3, 60% greater transmissibility and 80% immune escape.

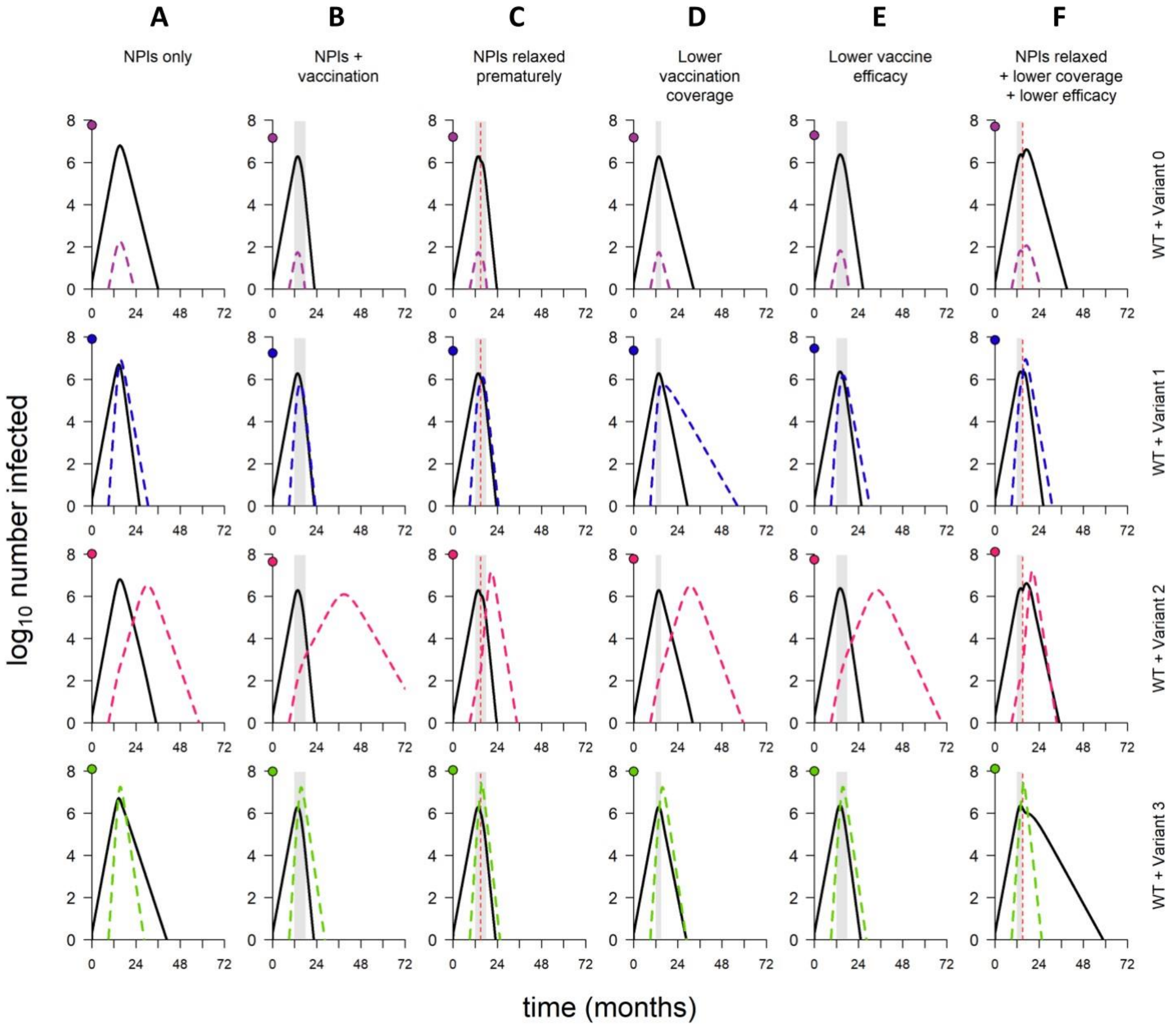


**Fig S11.** Dynamics of WT and variants, in simulations with varying combinations of control measures, for alternative model with higher degree of immune escape. (A) No vaccination but NPIs in place throughout. (B) Default model conditions (NPIs in place throughout, 100% vaccination coverage, 95% vaccine efficacy against WT). (C) NPIs lifted when vaccination coverage reaches 50% (other model conditions as in B). (D) 50% vaccination coverage (other model conditions as in B). (E) 70% vaccine efficacy (other model conditions as in B). (F) Combination of conditions C through E (NPIs relaxed, 50% vaccination coverage, and 70% vaccine efficacy). In each panel, point on y-axis indicates the total number of infections over the entire simulation. All simulations run for an extended duration of six years (default condition is three years); variant introduced at 9 months; in simulations with vaccination, rollout begins at 12 months and lasts 6 months if final coverage is 100%, 3 months if final coverage is 50%. Solid/black lines, WT; colored/dashed lines, variants; gray shading, vaccine rollout; dashed vertical line, 50% vaccination coverage. Variant phenotypes are as follows: variant 0, identical to WT; variant 1,

60% greater transmissibility; variant 2, 80% immune escape; variant 3, 60% greater transmissibility and 80% immune escape.

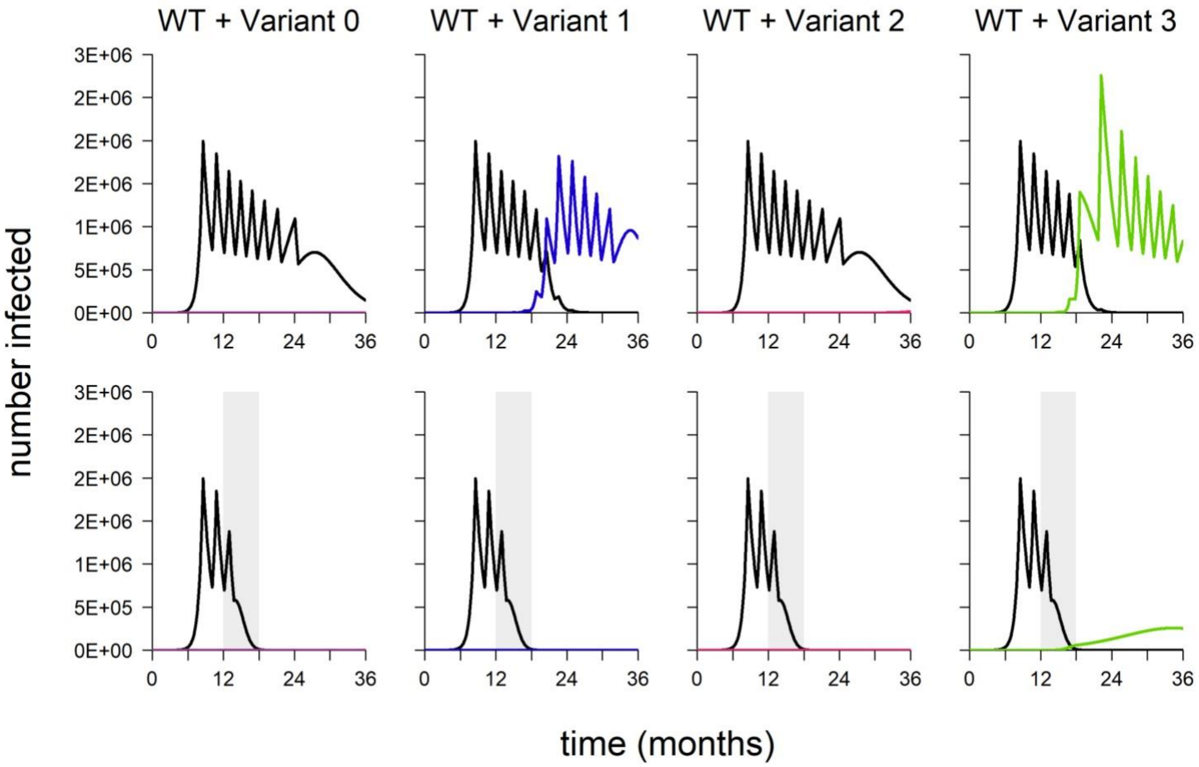


**Fig S12.** Dynamics of WT and variant strains under the alternative model with rolling lockdowns, without vaccination (top row) and with vaccination (bottom row). Black line, WT; colored line, variant; gray shading, vaccine rollout. Variant introduced at 9 months in all simulations; in simulations with vaccination, vaccine rollout begins at 12 months and lasts 6 months. Variant phenotypes are as follows: variant 0, identical to WT; variant 1, 60% greater transmissibility; variant 2, 40% immune escape; variant 3, 60% greater transmissibility and 40% immune escape.

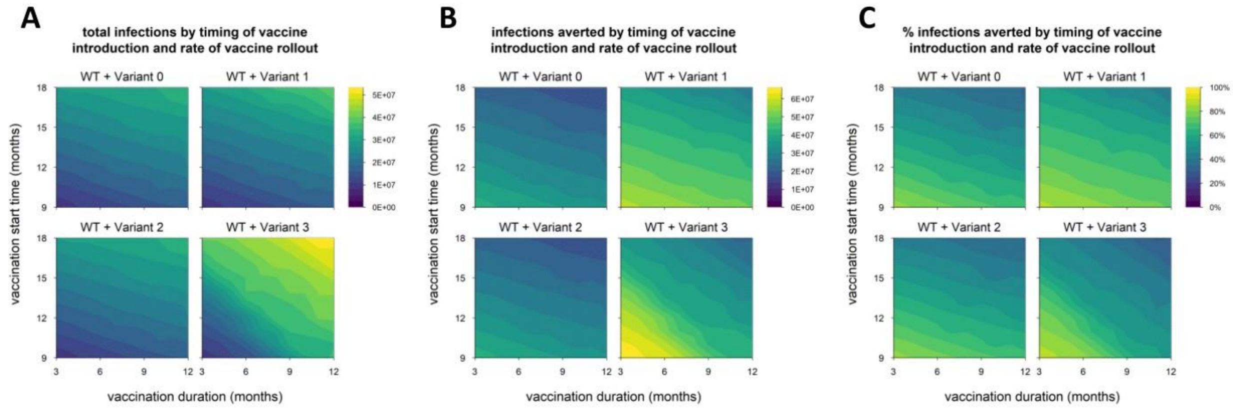


**Fig S13.** Epidemic size and vaccination impact for alternative model with rolling lockdowns. (A) Total infections (WT + variant) in simulations with each hypothetical variant, for varying rates of vaccination (vaccination duration, x-axis) and time of vaccine introduction (vaccination start time, y-axis); shaded contours represent total infections. (B) Number of infections (WT + variant) averted by vaccination; shading represents number of infections averted. (C) Percentage of infections (WT + variant) averted by vaccination; shading represents % infections averted. Variant introduced at 9 months in all simulations. Variant phenotypes are as follows: variant 0, identical to WT; variant 1, 60% greater transmissibility; variant 2, 40% immune escape; variant 3, 60% greater transmissibility and 40% immune escape.

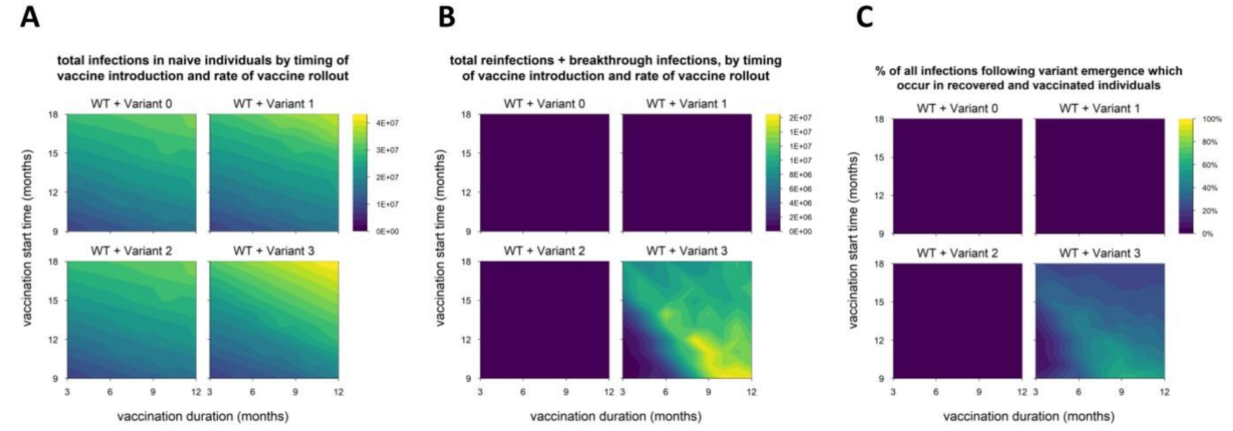


**Fig S14.** Breakdown of infections by immune status (naïve vs. recovered/vaccinated) for alternative model with rolling lockdowns. (A) Total infections in naïve individuals, in simulations with each variant, for varying rates of vaccination (x-axis) and time of vaccine introduction (y-axis); shaded contours represent total infections. (B) Total infections in recovered and vaccinated individuals (reinfections and breakthrough infections); shaded contours represent total infections. (C) Percentage of all infections occurring in recovered/vaccinated individuals (reinfections/breakthrough infections), starting from the time of variant emergence; shading represents percentage of infections occurring in recovered/vaccinated individuals. Variant introduced at 9 months in all simulations. Variant phenotypes are as follows: variant 0, identical to WT; variant 1, 60% greater transmissibility; variant 2, 40% immune escape; variant 3, 60% greater transmissibility and 40% immune escape.

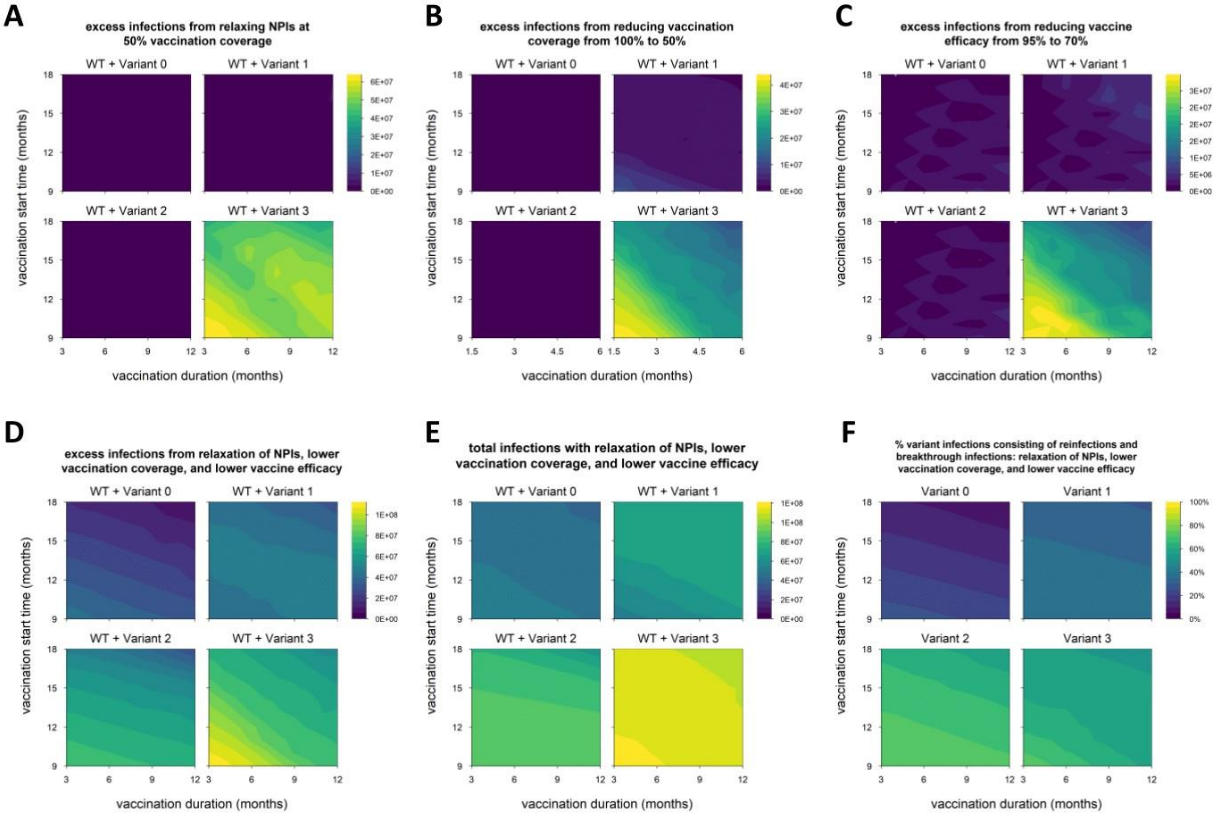


**Fig S15.** Impact of reduced control measures on epidemic size for alternative model with rolling lockdowns. (A) Excess infections from lifting nonpharmaceutical interventions (NPIs) when vaccination coverage reaches 50% (default condition is NPIs continued indefinitely); shading represents excess infections compared to default conditions/parameters. (B) Excess infections from reducing vaccination coverage from 100% to 50%. (C) Excess infections from reducing baseline vaccine efficacy (against WT) from 95% to 70%. (D) Excess infections from the combination of conditions A through C (lifting NPIs, reduced coverage and reduced efficacy). (E) Total infections (WT + variant) under the combined conditions of panels A through C; shading represent total infections. (F) Percentage of variant infections composed of reinfections and breakthrough infections under the combined conditions of panels A through C; shading represent percentage of variant infections occurring in recovered/vaccinated individuals. Variant introduced at 9 months in all simulations. Variant phenotypes are as follows: variant 0, identical to WT; variant 1, 60% greater transmissibility; variant 2, 40% immune escape; variant 3, 60% greater transmissibility and 40% immune escape.

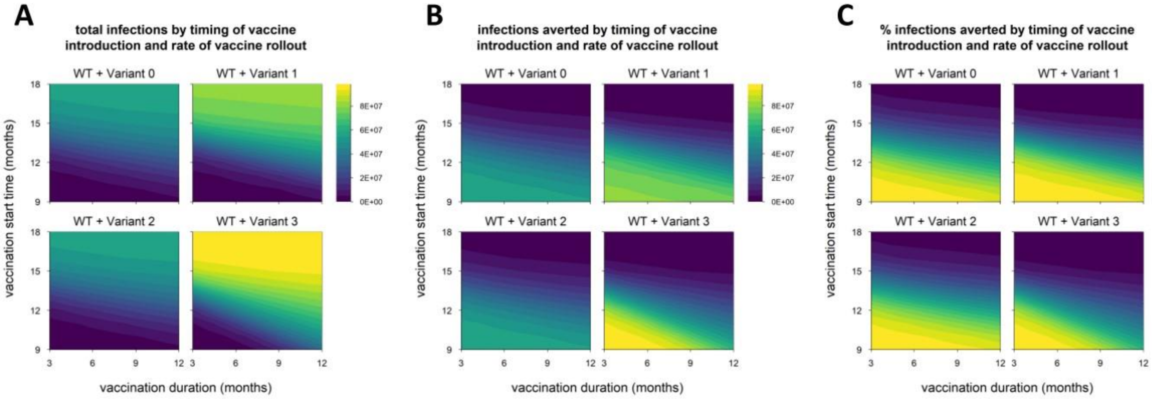


**Fig S16.** Epidemic size and vaccination impact for alternative model with all-or-nothing immunity. (A) Total infections (WT + variant) in simulations with each hypothetical variant, for varying rates of vaccination (vaccination duration, x-axis) and time of vaccine introduction (vaccination start time, y-axis); shaded contours represent total infections. (B) Number of infections (WT + variant) averted by vaccination; shading represents number of infections averted. (C) Percentage of infections (WT + variant) averted by vaccination; shading represents % infections averted. Variant introduced at 9 months in all simulations. Variant phenotypes are as follows: variant 0, identical to WT; variant 1, 60% greater transmissibility; variant 2, 40% immune escape; variant 3, 60% greater transmissibility and 40% immune escape.

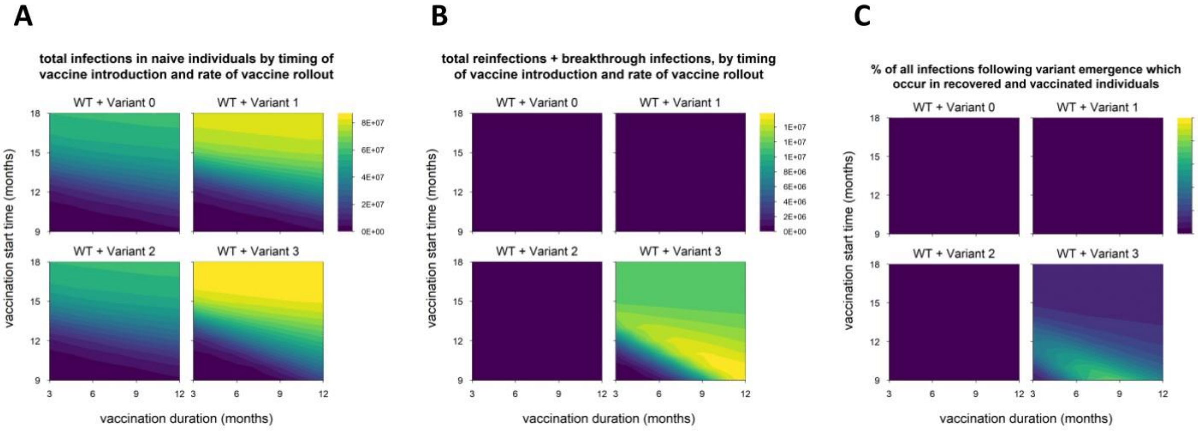


**Fig S17.** Breakdown of infections by immune status (naïve vs. recovered/vaccinated) for alternative model with all-or-nothing immunity. (A) Total infections in naïve individuals, in simulations with each variant, for varying rates of vaccination (x-axis) and time of vaccine introduction (y-axis); shaded contours represent total infections. (B) Total infections in recovered and vaccinated individuals (reinfections and breakthrough infections); shaded contours represent total infections. (C) Percentage of all infections occurring in recovered/vaccinated individuals (reinfections/breakthrough infections), starting from the time of variant emergence; shading represents percentage of infections occurring in recovered/vaccinated individuals. Variant introduced at 9 months in all simulations. Variant phenotypes are as follows: variant 0, identical to WT; variant 1, 60% greater transmissibility; variant 2, 40% immune escape; variant 3, 60% greater transmissibility and 40% immune escape.

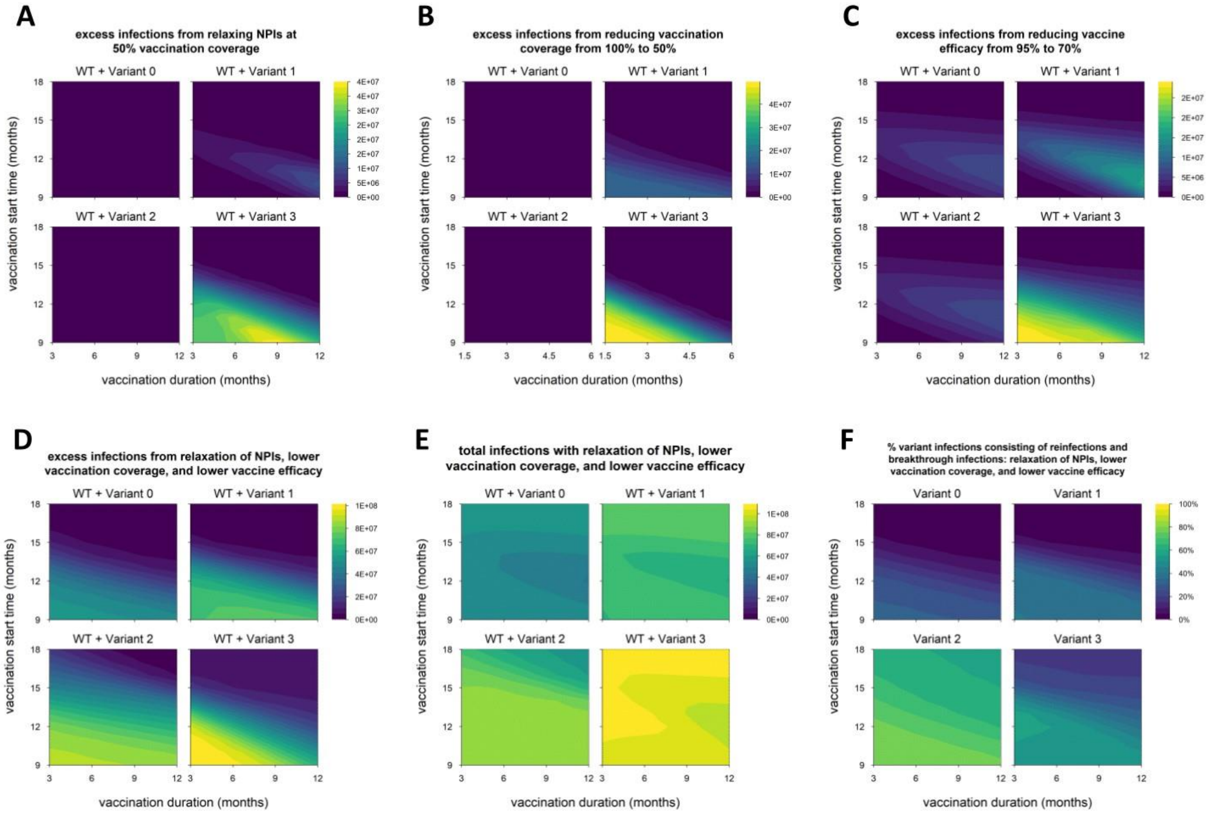


**Fig S18.** Impact of reduced control measures on epidemic size for alternative model with all-or-nothing immunity. (A) Excess infections from lifting nonpharmaceutical interventions (NPIs) when vaccination coverage reaches 50% (default condition is NPIs continued indefinitely); shading represents excess infections compared to default conditions/parameters. (B) Excess infections from reducing vaccination coverage from 100% to 50%. (C) Excess infections from reducing baseline vaccine efficacy (against WT) from 95% to 70%. (D) Excess infections from the combination of conditions A through C (lifting NPIs, reduced coverage and reduced efficacy). (E) Total infections (WT + variant) under the combined conditions of panels A through C; shading represent total infections. (F) Percentage of variant infections composed of reinfections and breakthrough infections under the combined conditions of panels A through C; shading represent percentage of variant infections occurring in recovered/vaccinated individuals. Variant introduced at 9 months in all simulations. Variant phenotypes are as follows: variant 0, identical to WT; variant 1, 60% greater transmissibility; variant 2, 40% immune escape; variant 3, 60% greater transmissibility and 40% immune escape.
